# SubGaitNet: A Decision-Oriented and Interpretable AI Framework for Robust GRF-Based Gait Assessment in Neurological and Musculoskeletal Care

**DOI:** 10.64898/2026.06.30.26356903

**Authors:** Wenhao Li, Shi Chang, Liujinxiang Zhu, Yihang Bao, Tianqi Liu, Han Wang, Guan Ning Lin

## Abstract

Ground reaction force (GRF)-based gait analysis provides objective, non-invasive evidence for neurological and musculoskeletal assessment, but its translation into medical AI decision support is limited by heterogeneous sensing devices, variable-length recordings, acquisition noise, sensor failures, and restricted access to high-cost gait laboratories. We propose SubGaitNet, a decision-oriented and interpretable AI framework designed to address four clinically relevant challenges in GRF-based medical AI: signal-length variability, sensing noise, long-range gait-phase dependency, and pathological frame-to-frame variability. SubGaitNet integrates GRF temporal slicing, multi-scale deep residual shrinkage, masked Transformer modeling, and a Sub-LSTM branch for adjacent-frame variability modeling. In subject-independent evaluation on two public clinical gait datasets, SubGaitNet achieved an AUC of 0.979 for Parkinson’s disease (PD) screening and an ACC of 0.940/F1-score of 0.910 for Hoehn & Yahr severity assessment using wearable pressure insoles. On the GaitRec force-plate dataset, SubGaitNet achieved ACC values of 0.951 and 0.918 for four-class and five-class musculoskeletal impairment assessment, respectively. Additional analyses showed stable bootstrap confidence intervals, calibrated PD screening probabilities (Brier score = 0.059; expected calibration error = 0.051), positive decision-curve net benefit across clinically relevant thresholds, and ordinally plausible H&Y errors. Robustness tests under simulated sensor failure, noise perturbation, and reduced-channel inputs supported the model’s stability under clinically plausible sensing uncertainty and accessibility constraints. SHAP explanations highlighted biomechanically meaningful hindfoot and forefoot regions. Overall, SubGaitNet provides a reusable, interpretable, and decision-support-oriented AI methodology for GRF-based gait health assessment, while prospective clinician-in-the-loop validation remains necessary before clinical deployment.

**Highlights:**

- SubGaitNet converts noisy and variable-length GRF recordings into clinically usable gait evidence.
- Its architecture targets four medical-AI failure modes: misalignment, noise, phase dependency, and pathological variability.
- A reusable framework supports PD screening, PD severity staging, and musculoskeletal impairment assessment.
- Calibration, decision-curve, and ordinal analyses assess outputs beyond discrimination accuracy.
- Stress testing and SHAP link robust predictions to heel-strike and push-off biomechanics.

## 1. Introduction

Gait reflects the coordinated function of the neural and musculoskeletal systems and provides clinically relevant information for evaluating motor impairment, disease progression, and rehabilitation status [1]. As a non-invasive and quantitative modality, ground reaction force (GRF)-based gait analysis has important potential for AI-supported medical assessment, including screening-oriented evaluation of neurodegenerative disorders such as Parkinson’s disease (PD), disease severity stratification, musculoskeletal impairment assessment, and longitudinal rehabilitation monitoring [2]. In clinical and community settings, GRF-related data can be acquired using heterogeneous sensing devices. Laboratory-grade force plates provide high-precision triaxial GRF signals and center-of-pressure (CoP) trajectories [3], whereas wearable pressure-sensor insoles capture distributed vertical plantar loading and are more suitable for continuous or home-based monitoring [4]. These sensing contexts differ substantially in signal semantics, channel configurations, acquisition environments, and data-quality profiles. Therefore, AI methods intended for medical gait assessment should not merely perform well on a single data format, but should remain reliable, interpretable, and clinically meaningful across representative gait-sensing conditions.

Traditional machine learning methods, including support vector machines (SVMs) [5] and k-nearest neighbors (KNN) [6], have been widely used for gait assessment, but their performance often depends on labor-intensive handcrafted features. For example, Shuzan et al. extracted 191 time- and frequency-domain features to classify healthy and impaired gait patterns [7]. Deep learning methods, including convolutional neural networks (CNNs), have reduced the dependence on manual feature design and enabled end-to-end learning from GRF signals. Representative examples include GaitRec-Net, which used a 1D-CNN architecture for gait disorder detection [8], and CNN-DWT variants that combine convolutional modeling with time-frequency analysis [9].

However, many existing methods remain designed around a specific device type, disease task, or sensing format. This limits their usefulness for medical decision support, where gait data may be collected using different devices and under imperfect acquisition conditions.

Several challenges are particularly important for medical AI based on GRF gait signals. First, heterogeneous sensing devices encode different physical information: force plates measure resultant foot-ground forces and CoP trajectories, whereas pressure-sensor insoles record distributed vertical loading at discrete plantar locations. Second, GRF recordings are variable in length because of differences in walking speed, acquisition duration, and patient motor state; naive padding or resampling may introduce artifacts that affect downstream inference. Third, clinically meaningful gait abnormalities are not always reflected by gross waveform morphology. In PD, for example, gait rhythm instability, inter-step variability, and altered heel-strike/toe-off dynamics may provide information relevant to screening and severity assessment [10]. Generic sequence models such as Transformers [11] and recurrent neural networks (RNNs) [12] can capture temporal dependencies, but may underrepresent subtle frame-to-frame pathological variability if not specifically designed for gait-related medical inference.

From the perspective of clinically meaningful medical AI, a useful GRF-based model should support decision-oriented medical tasks rather than simply apply a generic classifier to medical data. For gait assessment, this means that the model should handle variable-length physiological signals, remain robust to realistic sensing uncertainty, be reusable across clinically relevant gait-sensing contexts, and provide interpretable evidence for clinician review. These requirements motivate evaluation beyond predictive accuracy, including robustness testing, constrained-input assessment, probability reliability, decision-curve analysis, and explanation of learned decision evidence.

To address these challenges, we propose SubGaitNet, an interpretable and decision-oriented AI framework for GRF-based gait assessment. The central design principle is to decompose GRF gait modeling into four linked representation-learning problems: temporal alignment, noise-robust feature extraction, long-range gait-phase modeling, and adjacent-frame variability modeling. This clinically structured problem-to-module mapping is intended to distinguish SubGaitNet from a generic stack of sequence models and provides a testable rationale for ablation, robustness, and interpretability analyses.

We evaluate SubGaitNet in two representative medical gait-assessment contexts: wearable pressure-insole data for Parkinsonian gait screening and severity assessment, and force-plate data for musculoskeletal impairment assessment. These two contexts were selected to examine whether the same representation-learning design remains useful across neurological and musculoskeletal gait assessment, rather than to claim a single disease-agnostic diagnostic model. The intended output of the framework is auxiliary decision evidence for clinician review, not an autonomous diagnostic decision.

The main methodological and medical contributions of this work are summarized as follows:

- We propose SubGaitNet as a configurable framework for wearable pressure-sensor insole data and force-plate data. The aim is not to claim device-independent zero-shot transfer, but to provide a consistent AI methodology that can be adapted and retrained across clinically relevant gait-sensing formats without redesigning the backbone architecture.
- SubGaitNet organizes GRF modeling into temporal alignment, noise-robust feature extraction, long-range gait-phase modeling, and adjacent-frame variability modeling, thereby addressing key data-quality and temporal-structure challenges in medical gait AI.
- By incorporating adjacent-frame variability modeling, SubGaitNet is designed to capture rhythm instability and phase-transition irregularities that are relevant to screening-oriented and severity-oriented assessment, particularly in Parkinsonian gait.
- We evaluate SubGaitNet on two public clinical gait datasets using subject-independent protocols, covering PD screening, H&Y severity assessment, and musculoskeletal impairment classification. Beyond conventional metrics, the validation includes robustness testing, constrained-input assessment, probability calibration, decision-curve analysis, ordinal-aware severity analysis, and SHAP-based biomechanical interpretation.

## 2. Materials and methods

### 2.1 Datasets description

To evaluate SubGaitNet as an AI framework for GRF-based medical decision support, we employed two public clinical gait datasets: the Gait in Parkinson’s Disease dataset [13] and the GaitRec musculoskeletal impairment dataset [14]. These datasets represent two clinically relevant sensing scenarios: wearable pressure sensor insoles for continuous or community-based gait monitoring and force plates for standardized laboratory or rehabilitation-center assessment.

#### 2.1.1 Gait in Parkinson’s Disease dataset

To evaluate the performance of SubGaitNet in processing data from wearable pressure sensor insoles, we obtained the benchmark dataset "Gait in Parkinson’s Disease" from PhysioBank. This database comprises gait measurements from 93 patients with idiopathic PD and 73 healthy controls across three independent studies, yielding a total of 306 recordings. These three studies investigated the effects of dual tasks, Rhythmic Auditory Stimulation (RAS), and treadmill walking on gait, encompassing gait characteristics under diverse experimental conditions. The database also includes demographic information and disease severity graded by the Hoehn & Yahr (H&Y) scale. Vertical ground reaction forces were recorded as subjects walked on level ground at their self-selected, comfortable pace for approximately 2 minutes. Under each foot, eight sensors (Ultraflex Computer Dyno Graphy, Infotronic Inc.) were positioned to measure force variations over time (units: Newton). The outputs from these 16 sensors were digitally recorded at a sampling rate of 100 Hz.

All PD patients were clinically evaluated by neurologists and staged for disease severity according to the H&Y scale (mean score: 2.26 ± 0.34, covering mild to moderate severe stages). To preclude potential confounding effects of demographic factors on classifier performance, we conducted statistical significance tests between the two subject groups. As detailed in Supplementary Table 1, no statistically significant differences were observed between the PD group and the healthy control group regarding age (*p* = 0.066), height (*p* = 0.543) and weight (*p* = 0.774). This procedure suggests that the features learned by the model originate primarily from pathological alterations in gait dynamics, rather than biases induced by body size or age.

#### 2.1.2 GaitRec dataset

As a complement to the wearable data, we selected the large-scale GaitRec dataset to evaluate the model on force plate data. This dataset comprises complete gait records from 2,084 patients with diverse musculoskeletal functional impairments and 211 healthy controls. All data were manually annotated by senior physical therapists with over ten years of clinical experience. Based on the patients’ original medical diagnostic reports, the samples were initially categorized into two primary classes: Control (healthy) and Impaired (gait disorder). Subsequently, the Impaired category was further subdivided into four classes according to the anatomical site of the primary injury: Hip (H), Knee (K), Ankle (A), and Calcaneus (C).

Data acquisition was conducted in a standard gait analysis laboratory, where subjects were instructed to walk along a level walkway at their self-selected, comfortable pace. Two force plates (Kistler, Type 9281B12, Winterthur, CH) embedded in the walkway were utilized to synchronously record the kinetic signals of bilateral feet. For each walking trial, the dataset provides ground reaction forces in three orthogonal directions, along with the trajectory coordinates of the CoP. As shown in Supplementary Table 2, the subject cohort spans a broad range of ages and body types, encompassing individuals from young adults to the elderly across various stages of rehabilitation. Regarding class distribution, data for knee injuries (625 individuals, 19,873 trials) and ankle injuries (627 individuals, 21,386 trials) are the most abundant, whereas calcaneus injuries are relatively scarce. This inter-class data imbalance reflects the actual distribution of clinical visits and may impose higher demands on the robustness of the classification model.

### 2.2 SubGaitNet framework for GRF-based medical decision support

In real-world clinical and healthcare applications, GRF-based gait assessment must handle device heterogeneity, variable-length recordings, noisy acquisition, and incomplete sensor information. To address these issues, we propose SubGaitNet, a robust and interpretable AI framework designed to support screening-oriented and assessment-oriented medical tasks from heterogeneous GRF signals. The overall architecture contains four components: (i) the GRF Temporal Slicing module for structured signal alignment; (ii) the MS-DRSN for noise-robust feature extraction; (iii) the Dual-branch Temporal Modeling module, including a masked Transformer encoder and a Sub-LSTM branch; and (iv) the downstream task classifier for clinically relevant gait assessment.

To resolve the curse of dimensionality and temporal alignment issues caused by variable-length signals, we design the GRF Temporal Slicing module. It partitions the raw GRF signals into a sequence of fixed-length GRF frames, with each frame representing a localized phase within the gait cycle (e.g., the instant of heel-strike or toe-off). Consequently, continuous signals are transformed into structured GRF frame sequences. Subsequently, these frames are fed in parallel to the MS-DRSN module. This module leverages convolutional kernels of varying sizes to capture multi-scale temporal patterns, while simultaneously utilizing an adaptive soft-thresholding mechanism [14] to filter signal noise, thereby significantly enhancing feature robustness.

After extracting robust temporal feature sequences, the model proceeds to the Dual-branch Temporal Modeling stage. The first branch employs a Transformer encoder, utilizing the multi-head attention mechanism to capture long-range dependencies and phase transition patterns between GRF frames. We incorporate a padding mask to ignore zero-padded positions introduced for alignment, effectively mitigating information interference during variable-length sequence processing. The parallel second branch, the Sub-LSTM, is dedicated to extracting gait variability features. Specifically, the Sub-LSTM models the difference sequences between adjacent GRF frames, extracting the dynamic transition from one step to the next. This design enables the model to capture the gait rhythm associated with neurological disorders such as PD. Ultimately, the features derived from both branches are forwarded to the classification module. The overall architecture of SubGaitNet is illustrated in Fig. 1.

**Fig. 1.**
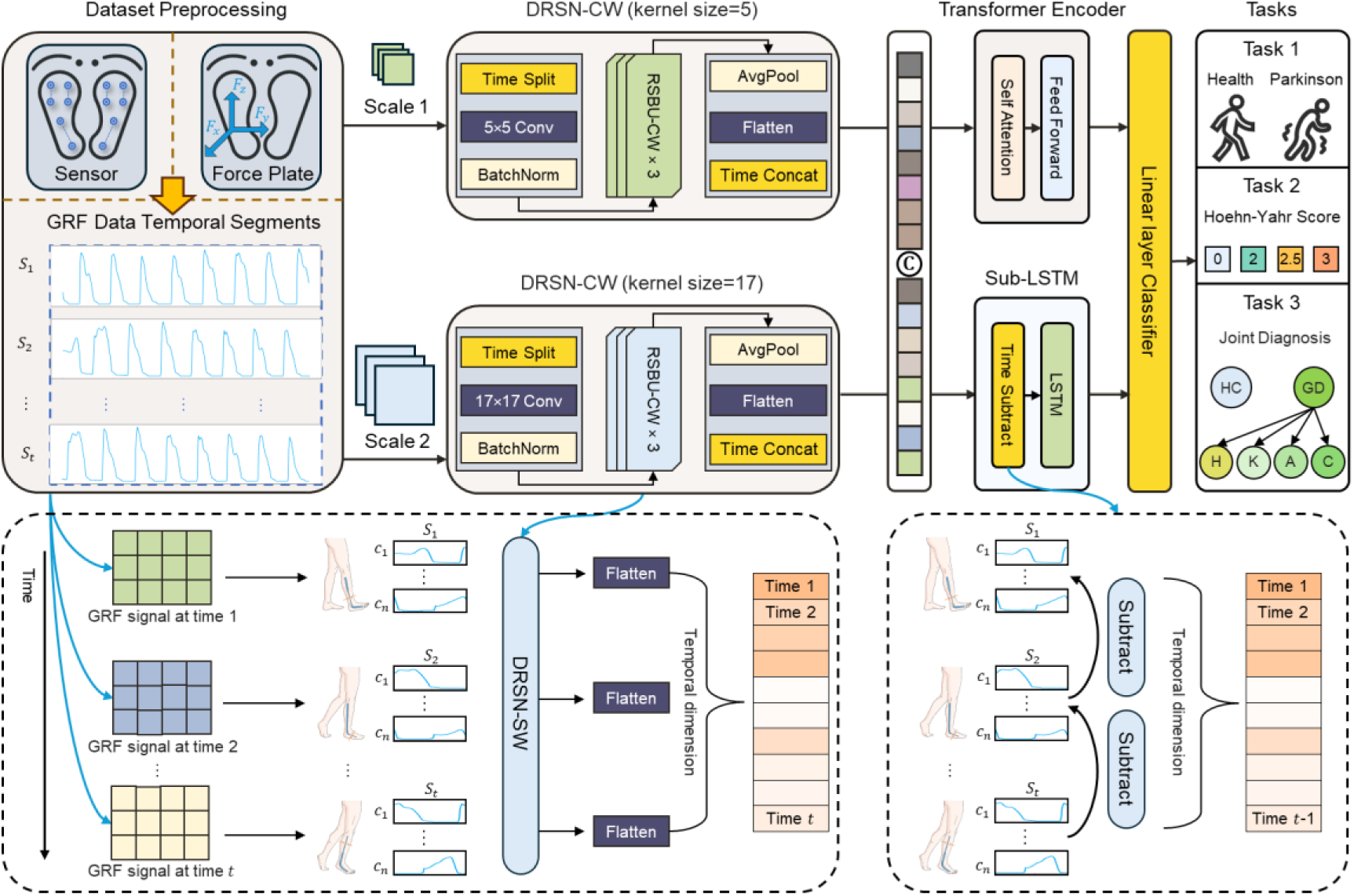
Framework of the proposed SubGaitNet model.

Although force plates and pressure sensor insoles measure different physical quantities, both can be represented as multivariate time series after structured temporal slicing. Based on this shared representation, SubGaitNet retains the same core architecture across both data types: multi-scale temporal convolutions extract local kinetic patterns, the Transformer encoder models long-range phase dependencies, and the Sub-LSTM branch captures gait variability. This shared architecture should be interpreted as evidence for methodological reusability across representative clinical gait sensing contexts, not as evidence of direct zero-shot transfer between devices. Device-specific retraining and calibration remain necessary in the current study, and this constraint is explicitly considered when interpreting the results .

### 2.3 GRF temporal slicing module

To model the gait cycle information in GRF signals, we propose the GRF Temporal Slicing module, which partitions the raw signals into structured GRF frame sequences. In this study, the GRF dataset is defined as *X* = {*X*_1_, *X*_2_, …, *X_N_*}, where *N* denotes the total number of samples. Each sample comprises *C* channels (representing different sensors or varying GRF directions), formulated as 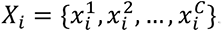. The signal for each channel is denoted as 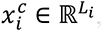, where *L_i_* represents the temporal length of the *i*-th sample. In real-world gait data, signal lengths frequently vary across samples due to discrepancies in acquisition duration and individual patient gait characteristics. To facilitate unified model processing, all signals are first padded with zeros to a uniform length *L* via a zero-padding operation. Subsequently, the length-aligned GRF signal is divided into *T* non-overlapping frames, represented as 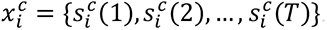. Here, the number of frames is *T* = ⌊*L*/*L_s_*⌋, *L_s_* is the predetermined frame length 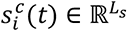, and denotes the *t*-th GRF frame. Fig. 2 illustrates the procedure for partitioning GRF temporal slices from raw signals.

**Fig. 2.**
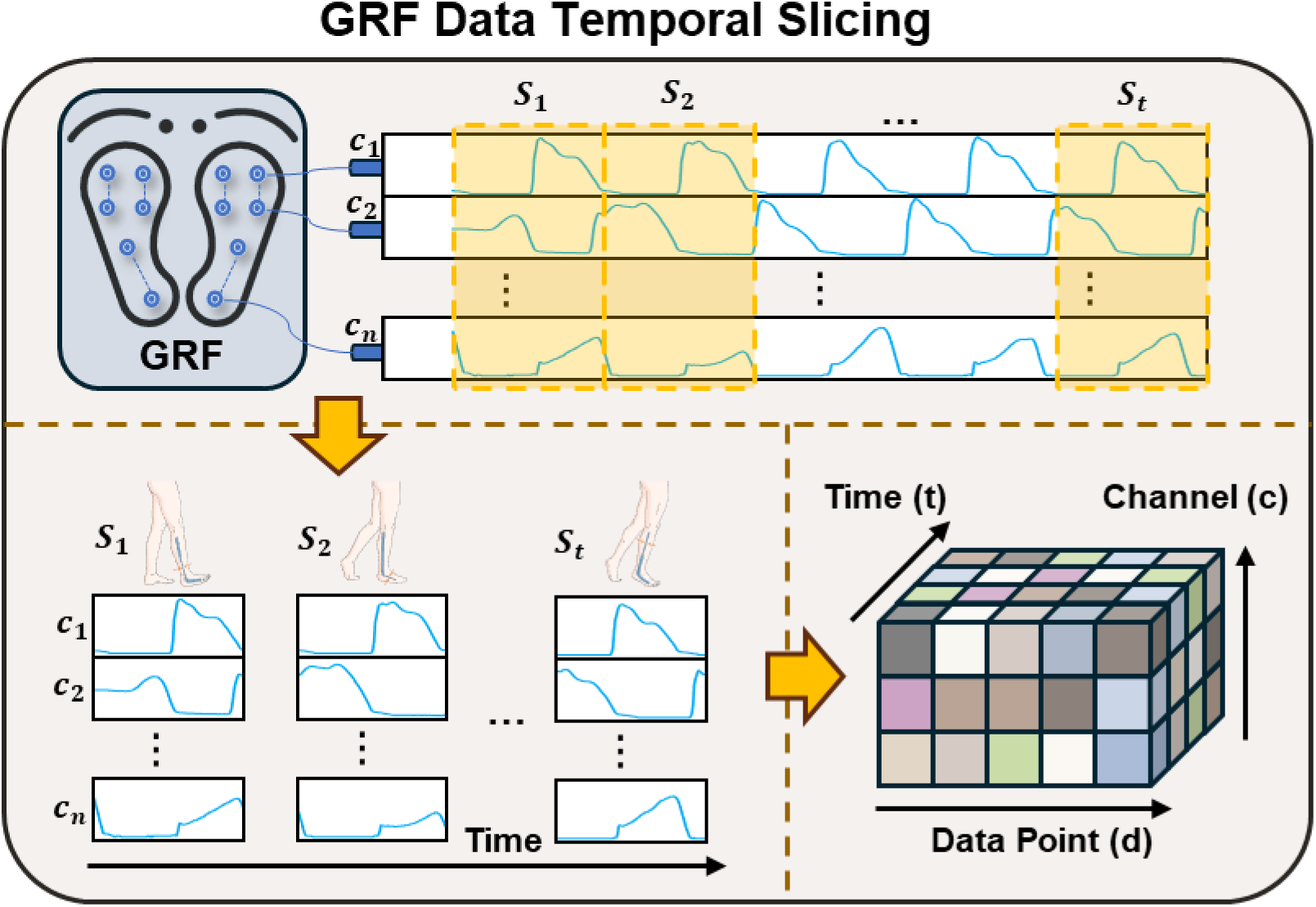
GRF Temporal Slicing module.

The rationale for temporal slicing is motivated by both biomechanical and computational perspectives. First, from a biomechanical standpoint, the partitioning of temporal slices corresponds to specific phases within the gait cycle, enabling each GRF frame to focus on extracting localized features of that particular phase. By sequentially concatenating multiple GRF frames, the model preserves the overarching periodic patterns of the GRF signals while capturing the dynamic transitions between phases, thereby revealing gait mechanisms more accurately. Second, computationally, temporal slicing helps mitigate the curse of dimensionality associated with processing exceedingly long signals. Furthermore, by configuring the partitioning granularity, a balance can be struck between preserving signal details and mitigating computational overhead. Shorter GRF frames are more effective for capturing the localized features of the signal, whereas longer slices are better suited for accentuating periodic patterns. The temporal slicing parameters utilized for the Gait in Parkinson’s Disease and the GaitRec datasets are detailed in Supplementary Table 3.

### 2.4 Multi-scale deep residual shrinkage module

GRF signals possess complex temporal features and multi-scale dynamic patterns. However, noise perturbations during the acquisition can degrade the quality of their feature representation. To extract features from GRF frames more effectively, we design a Multi-Scale Deep Residual Shrinkage Network (MS-DRSN). By integrating feature patterns across different receptive fields and incorporating a channel-wise adaptive soft-thresholding mechanism, this module aims to improve the model’s ability to model complex sequential signals.

The MS-DRSN comprises two parallel Deep Residual Shrinkage Network branches with Channel-wise thresholds (DRSN-CW) [15]. The first branch utilizes 5×5 convolutional kernels to extract fine-grained features of local temporal segments, whereas the second branch employs 17×17 convolutional kernels to capture broader dynamic patterns over longer temporal contexts. Each DRSN-CW branch is based on the ResNet-18 [16] framework, wherein the standard residual blocks are replaced by stacked Residual Shrinkage Building Units with Channel-wise thresholds (RSBU-CW). This substitution enhances the noise robustness of the extracted features. The architecture of the DRSN-CW is illustrated in Fig. 3.

**Fig. 3.**
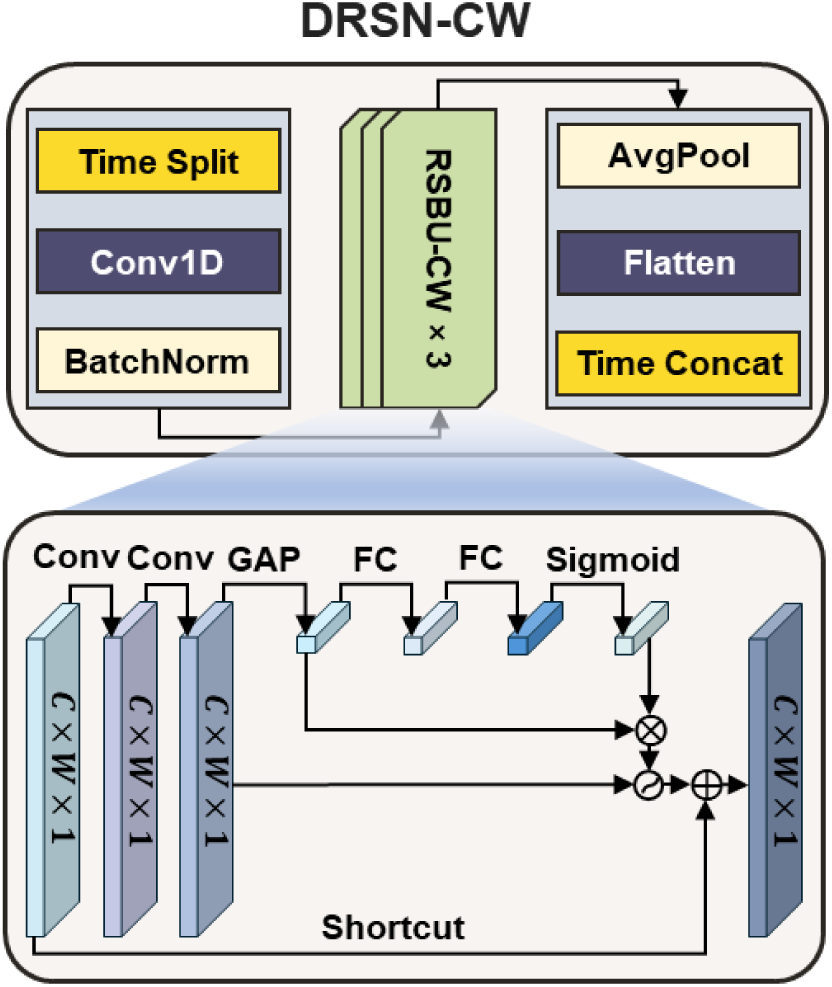
The architecture of DRSN-CW.

The RSBU-CW module serves as the fundamental building block of the DRSN-CW. Equipped with the soft-thresholding mechanism, its primary function is to suppress redundant features and emphasize salient features via adaptively computed channel-wise thresholds. Specifically, the input GRF frame *s* is first compressed into a one-dimensional vector through a Global Average Pooling (GAP) operation. Subsequently, it is processed by a two-layer Fully Connected (FC) network to compute the scaling parameter *α_c_*:

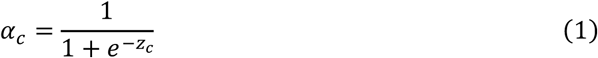

where *z_c_* represents the output feature value corresponding to the *c* -th channel from the FC network. Subsequently, the channel-wise threshold *τ_c_* is computed using (2):

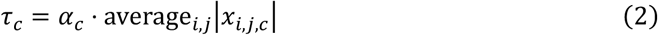

where average*_i_*_,*j*_ denotes the absolute average value of the feature map along *c*-th channel, and *i* and *j* are the spatial indices for width and height, respectively. Utilizing this dynamic threshold, a soft-thresholding shrinkage operation is executed at each spatial position of the input *s*, defined as follows:

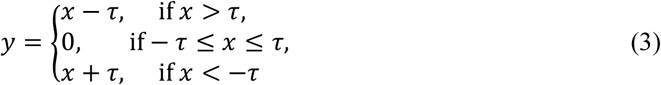

where *y* represents the output feature after shrinkage. This design facilitates flexible noise suppression across channels, thereby augmenting the model’s capacity to capture salient features.

Within each DRSN-CW, the RSBU-CW modules are cascaded via a residual connection strategy. This ensures that signal features are effectively preserved during propagation and significantly accelerates the network optimization process. For a given residual unit, the feature propagation is formulated as:

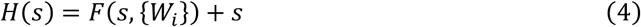

where *H*(*s*) denotes the output of the residual unit, and *F*(*s*, {*W_i_* }) represents the feature transformation through the RSBU-CW module. Finally, the two DRSN-CW branches with distinct convolutional kernel sizes yield output features *F_small_* (*x*) and *F_large_* (*x*), respectively. These multi-scale features are subsequently integrated via a concatenation operation, as formalized in the following equation:

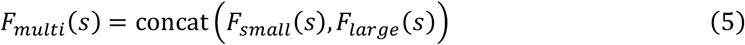

### 2.5 Dual-branch temporal modeling module

#### 2.5.1 Temporal Transformer encoder block

Following feature extraction via the MS-DRSN, the feature vectors of all GRF frames are concatenated chronologically to generate a unified time series representation. The length of this sequence equals the total number of GRF frames. This sequence serves as the input to the Transformer encoder, which is designed to model dynamic dependencies between gait phases and transition patterns.

The Transformer encoder comprises stacked multi-head self-attention layers and feed-forward neural networks. Its core component, the multi-head self-attention layer, extracts global dependencies within the time series through multiple scaled dot-product attention operations. As illustrated in Fig. 4, the inputs to the self-attention layer consist of queries *Q*, keys *K*, and values *V*. The attention function is computed as follows:

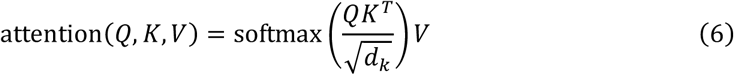

**Fig. 4.**
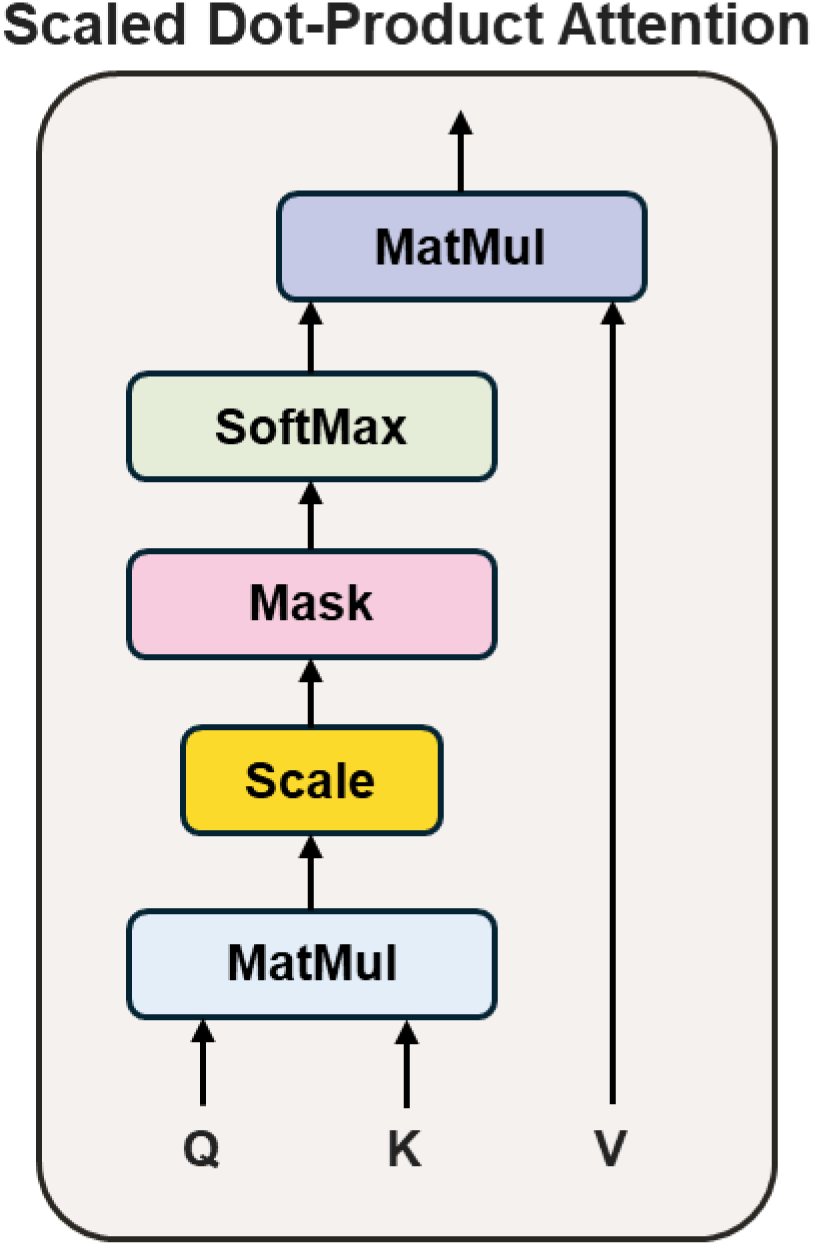
Scaled dot-product attention.

where *d_k_* represents a scaling factor determined by the dimension of the keys, and the matrices *Q*,

*K*, and *V* are derived from the input sequence features via distinct linear transformations. This mechanism enables the Transformer encoder to learn the weighted relationships across different GRF frames, thereby effectively capturing temporal dependencies. Furthermore, to address the invalid GRF frames introduced by zero-padding for temporal alignment, we apply a padding mask. This mask excludes padded positions from attention computation, reducing spurious effects on subsequent feature learning, thereby improving the model’s ability to extract salient signal features.

#### 2.5.2 Subtraction Long Short-Term Memory block

In parallel with the Transformer branch, which captures global dependencies, we design the Sub-LSTM module to explicitly model gait variability. The core pathological signature of neurodegenerative diseases, particularly PD, frequently manifests not merely as abnormal plantar pressure patterns, but more critically as rhythmic instability between adjacent gait cycles. To this end, this module initially performs a subtraction operation on the time-series output from the MS-DRSN. Specifically, for adjacent GRF frames *t* and *t* − 1, the difference between their feature vectors is calculated as Δ*x_t_* = *x_t_* − *x_t_*_−1_.

The generated differential feature sequence is subsequently fed into a Long Short-Term Memory (LSTM) network [17]. The LSTM is a specialized RNN that effectively mitigates the vanishing gradient problem inherent in long-sequence training by incorporating gating mechanisms. As illustrated in Fig. 5, the LSTM unit regulates the information flow via a forget gate, an input gate, and an output gate. Its core computation process is formulated as follows:

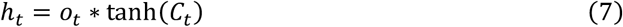

**Fig. 5.**
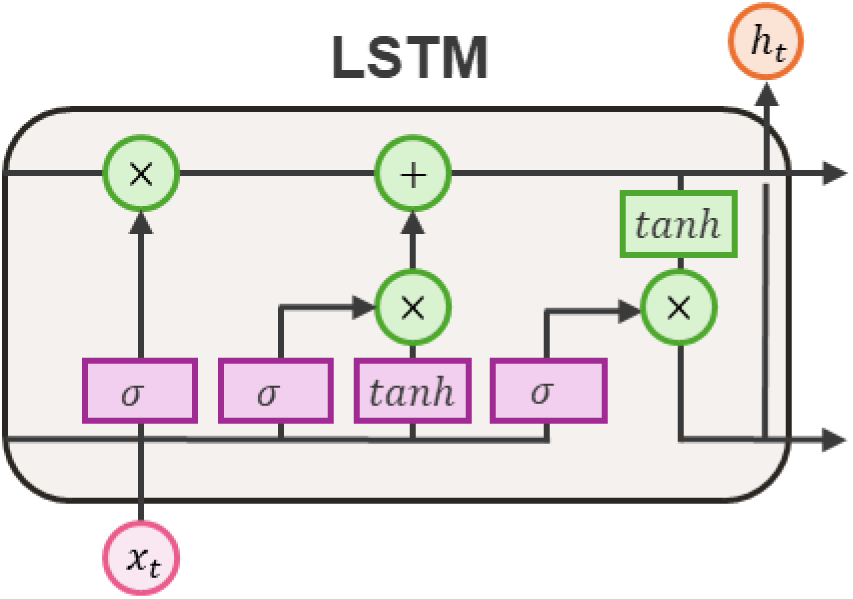
The architecture of Long Short-Term Memory.

where *o_t_* represents the output gate, ∗ denotes element-wise multiplication, *C_t_* and ℎ*_t_* signify the cell state and hidden state at time *t*, respectively. Through this mechanism, the Sub-LSTM can selectively discard irrelevant random fluctuations while accumulating pathological variability patterns that possess diagnostic value.

Ultimately, the final hidden state output by the Sub-LSTM branch is considered the representation of gait variability. To achieve a comprehensive characterization of gait, we concatenate the global temporal features extracted by the Transformer branch with the differential features derived from the Sub-LSTM. The concatenated features are subsequently fed into a fully connected layer and a Softmax classifier for final classification. Model parameters are optimized using the cross-entropy loss, which is defined as follows:

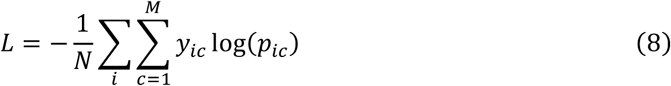

where *M* denotes the total number of classes, and *y_ic_* acts as a binary indicator (taking a value of 0 or 1) such that *y_ic_* = 1 if the ground-truth class of sample *i* is *c*, and 0 otherwise. Additionally, *p_ic_* represents the predicted probability that the observed sample *i* belongs to class *c*.

### 2.6 Implementation and training details

In this study, we evaluated the proposed model on two public datasets: the Gait in Parkinson’s Disease dataset and the GaitRec dataset. The Gait in Parkinson’s Disease dataset primarily includes gait recordings from patients with PD. On this dataset, we conducted two distinct classification tasks: (i) a binary classification task distinguishing PD patients from healthy controls (HC), and (ii) a four-class classification task assessing the severity stages of PD. Additionally, the GaitRec dataset comprises gait data from healthy individuals and subjects with musculoskeletal impairments. On the GaitRec dataset, we performed a four-class classification task (A, C, H, K) and a five-class classification task (HC, A, C, H, K).

The model was implemented in PyTorch 2.1.0 and trained on a single NVIDIA RTX 3090 GPU. The learning rate was set to 1 × 10^−5^, and the Adam optimizer was employed for parameter updates. The training process spanned a total of 100 epochs, with the batch size configured to 32. Regarding the MS-DRSN module, the kernel sizes for the small and large convolutional branches were set to 5 × 5 and 17 × 17, respectively. The Transformer encoder consisted of two layers, with each layer equipped with two attention heads. The Sub-LSTM module was composed of a 2-layer LSTM network, where the hidden sizes for the first and second layers were 512 and 256, respectively. Detailed parameter configurations are summarized in Table 1.

**Table 1.** List of hyperparameters.

| Hyperparameter | Value |
| --- | --- |
| Learning rate | $1 \times 10^{-5}$ |
| Weight decay | $1 \times 10^{-4}$ |
| Batch size | 32 |
| Epoch | 100 |
| Small conv size | 5 |
| Large conv size | 17 |
| Number of attention heads | 2 |
| Number of encoder layers | 2 |
| Hidden size of encoder | 2048 |
| Number of LSTM layers | 2 |
| Hidden size of the first LSTM | 512 |
| Hidden size of the second LSTM | 256 |

### 2.7 Medical decision-oriented evaluation protocol

For Medical AI, evaluation should demonstrate not only discrimination performance but also the potential usefulness of the AI method for decision-based clinical tasks. We therefore designed the evaluation protocol around four clinically relevant questions: (i) whether SubGaitNet can support screening-oriented and severity-oriented gait assessment across heterogeneous acquisition devices; (ii) whether the model remains stable under noisy or incomplete wearable measurements; (iii) whether potentially useful performance can be maintained under reduced-channel inputs that may enable lower-cost assessment in community or rehabilitation settings; and (iv) whether the model provides interpretable evidence aligned with biomechanically plausible gait regions.

This evaluation design was chosen to reflect how a medical AI model for gait assessment may be used in practice: a model output should be statistically reliable, probabilistically interpretable, robust to sensing uncertainty, and explainable enough to support clinician review rather than to replace clinical judgment.

To evaluate applicability across clinical sensing contexts, we selected wearable pressure sensor insole data and laboratory force-plate data. The former captures plantar pressure distribution and is suitable for continuous or home-based monitoring, whereas the latter measures whole-body GRF and CoP trajectories and is commonly used in standardized gait laboratories. The backbone architecture of SubGaitNet was kept invariant, with only input-dimension-specific adaptation and dataset-specific retraining. This design does not assume direct model transferability between devices; instead, it tests whether the same AI framework can be reused across different health-care gait acquisition formats while supporting distinct decision-oriented tasks.

For robustness evaluation, we focused on wearable pressure sensor insoles because these data are particularly susceptible to imperfect acquisition during long-term or non-laboratory monitoring. We simulated two clinically relevant data-quality problems: noise perturbation and sensor failure. The noise perturbation experiment superimposed random noise onto raw signals to assess performance under degraded signal quality. The sensor failure experiment zeroed out input channels to simulate poor sensor contact, hardware aging, or local sensor malfunction. These analyses were designed to test whether model outputs remain reliable under realistic sensing uncertainty.

For constrained-input evaluation, we used the GaitRec force-plate dataset to simulate simplified sensing configurations. We compared full 3D GRF plus CoP inputs with force-only and vertical-GRF-only inputs. This experiment was designed to assess whether a reduced sensing configuration could still provide useful decision-support information, which is important for accessible gait assessment in lower-resource or community settings.

Finally, because AI systems in medicine require trustworthiness and clinically understandable evidence, we used SHapley Additive exPlanations (SHAP) to examine whether model attributions were concentrated in biomechanically meaningful gait regions. The purpose was not to claim mechanistic causality, but to assess whether the learned decision evidence aligns with known gait-phase and plantar-loading patterns relevant to neurological impairment.

### 2.8 Baseline methods and evaluation metrics

To comprehensively validate the structural applicability and robustness of SubGaitNet in processing heterogeneous gait data, particularly its adaptability to laboratory force plate data (GaitRec) and wearable sensor data (Gait in Parkinson’s Disease), we selected two representative categories of methods for comparative evaluation. For the force plate data, we selected GaitRec-Net [18] as the deep learning baseline, a model explicitly designed for the multi-channel features of 3D GRF signals. Concurrently, we incorporated CNN-DWT [9], which integrates time-frequency analysis, along with KNN [7] and DWT [6] methods based on handcrafted statistical features.

For the pressure sensor insole data, we selected methods emphasizing sequential modeling, including 1D-Convnet [19] for capturing local morphological features and Uni-LSTM [20] for modeling long-term dependencies. To benchmark state-of-the-art temporal modeling capabilities, we employed 1D-Transformer [21] as a baseline. Furthermore, nonlinear dynamic methods [22] based on Phase Space Reconstruction (PSR) and Empirical Mode Decomposition (EMD) were included to serve as machine learning baselines. To fairly assess the models’ adaptability across heterogeneous gait data, we adopted a unified validation strategy. Regardless of whether the comparative methods were originally designed for force plates or pressure insoles, all models (including SubGaitNet) were independently trained and tested on both datasets.

To ensure the robustness of the evaluation, we split the training, validation, and testing sets across all datasets in an 8:1:1 ratio on a subject-independent basis. This strategy reduces the risk of artificially inflated performance caused by subject-level data leakage. The primary discrimination metrics included Accuracy (ACC), Precision, Recall, F1-score, Area Under the Receiver Operating Characteristic Curve (AUC), and Area Under the Precision-Recall Curve (AUPRC). For multi-class classification tasks, macro-averaging was used to balance the weights across classes. The macro-averaging is formulated as follows:

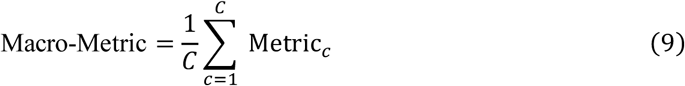

where *C* represents the total number of classes, and Metric*_c_* denotes the metric value computed independently for each class *c* . All relevant hyperparameters and training configurations are provided comprehensively in the code repository to support the reproducibility of the results.

### 2.9 Statistical uncertainty, calibration, and clinical utility analyses

To further strengthen the statistical reliability of model evaluation and to better align the experimental validation with the requirements of medical artificial intelligence, we performed additional analyses of statistical uncertainty, probability calibration, and potential clinical utility. All supplemental analyses were conducted using the same test splits and trained model weights as those used in the primary experiments. No additional data splitting, model retraining, or parameter updating was performed, thereby ensuring consistency and comparability with the main experimental results.

First, to quantify the uncertainty of performance estimates, bootstrap resampling was used to compute 95% confidence intervals for the primary evaluation metrics. For each main task, including Parkinson’s disease screening, Hoehn & Yahr (H&Y) severity grading, four-class musculoskeletal impairment assessment on GaitRec, and five-class musculoskeletal impairment assessment on GaitRec, test samples were resampled with replacement. Accuracy, macro-F1, AUC, and AUPRC were recalculated for each bootstrap sample. For multi-class tasks, AUC and AUPRC were computed using a one-vs-rest strategy with macro-averaging. The 2.5th and 97.5th percentiles of the bootstrap distributions were reported as the 95% confidence intervals.

Second, probability calibration analysis was performed for the binary Parkinson’s disease screening task. Because the output probability in this screening-oriented task can be interpreted as the estimated likelihood of Parkinsonian gait abnormality, we evaluated the original model probabilities without fitting any additional calibration model on the test set. Calibration curves were generated to compare the mean predicted probability with the observed positive fraction across probability bins. In addition, the Brier score was used to quantify the overall error of probabilistic prediction, and the expected calibration error (ECE) was calculated to measure the weighted average discrepancy between predicted probabilities and observed event frequencies.

Third, decision curve analysis (DCA) was conducted for the Parkinson’s disease screening task to evaluate the potential decision-related clinical utility of the model. Across a range of threshold probabilities, the net benefit of SubGaitNet was calculated and compared with two reference strategies: treating all subjects as positive and treating none as positive. This analysis was designed to examine whether the predicted probabilities could provide additional net benefit for screening-oriented clinical decision-making under different risk-threshold assumptions.

Finally, because H&Y staging has an inherent ordinal structure, ordinal-aware evaluation metrics were added for the Parkinson’s disease severity grading task. In addition to conventional multi-class discrimination metrics, we calculated the quadratic weighted kappa, adjacent-stage error rate, non-adjacent-stage error rate, and mean absolute H&Y stage error. These analyses were used to determine whether misclassifications mainly occurred between clinically adjacent stages and whether the model captured the continuous nature of disease severity progression. Compared with standard nominal multi-class metrics, these ordinal-aware measures provide a more clinically meaningful evaluation of the H&Y severity assessment task.

## 3. Results

### 3.1 Wearable insole-based Parkinsonian gait assessment under sensing uncertainty

We first evaluated SubGaitNet in a wearable pressure sensor insole scenario using the Gait in Parkinson’s Disease dataset. The tasks included screening-oriented discrimination between PD and healthy controls and fine-grained H&Y severity assessment. Because wearable sensing is particularly relevant to community, home, and longitudinal monitoring, we further evaluated model reliability under sensor failure and noise perturbation. These experiments assess whether SubGaitNet can provide robust gait-based decision support under realistic data-quality variation.

#### 3.1.1 Screening-oriented assessment of Parkinsonian gait

We first evaluated SubGaitNet on pressure sensor insole data for screening-oriented Parkinsonian gait assessment. As illustrated by the confusion matrix in Fig. 6a, the model yielded three misclassifications (one false positive and two false negatives) on the test set comprising 61 subjects. The corresponding receiver operating characteristic (ROC) curve (Fig. 6b) further supported this result, with an AUC of 0.979. These results suggest that SubGaitNet can extract discriminative gait patterns from wearable pressure signals and may provide auxiliary evidence for screening-oriented assessment; however, such outputs should not be interpreted as an autonomous diagnostic decision or a replacement for clinician evaluation.

**Fig. 6.**
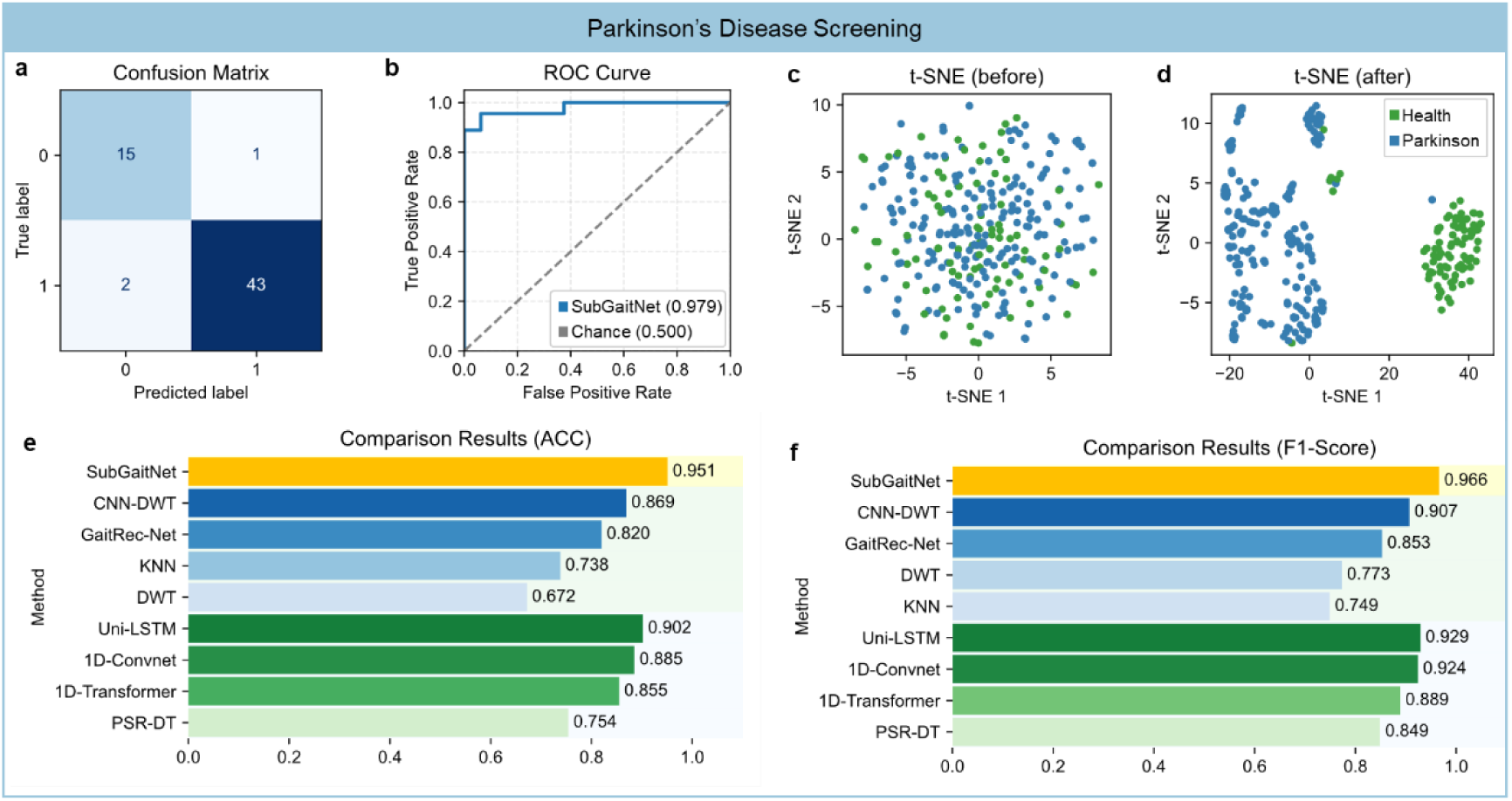
Performance of SubGaitNet for PD screening on the Gait in Parkinson’s Disease dataset.

To examine the learned representation, we used t-SNE to visualize feature distributions before and after network processing. As shown in Fig. 6c, healthy controls and PD samples overlapped substantially in the raw feature space, reflecting the nonlinear structure of the original GRF signals. After SubGaitNet-based feature extraction and temporal encoding (Fig. 6d), the two groups showed improved visual separation in the learned feature space. This visualization provides supportive, but not definitive, evidence that the model learned discriminative gait representations.

We compared SubGaitNet with two groups of baseline methods; the detailed results are summarized in Table 2. As illustrated in Fig. 6e-f, SubGaitNet outperformed all baseline models in terms of accuracy (0.951) and F1-score (0.966). Notably, the comparison shows a clear trend: methods designed for force plates (blue) exhibited lower overall performance on this dataset compared to those designed for pressure sensor insoles (green). For instance, the ACC of GaitRec-Net (0.820) was substantially lower than that of Uni-LSTM (0.902). This performance gap suggests that force plate-oriented methods struggle to effectively extract discriminative features from pressure sensor insole data. However, even the best-performing method designed for pressure sensor insoles (Uni-LSTM) fell behind SubGaitNet. This demonstrates that SubGaitNet can accurately characterize the plantar pressure distribution patterns inherent in pressure sensor insole data.

**Table 2.**
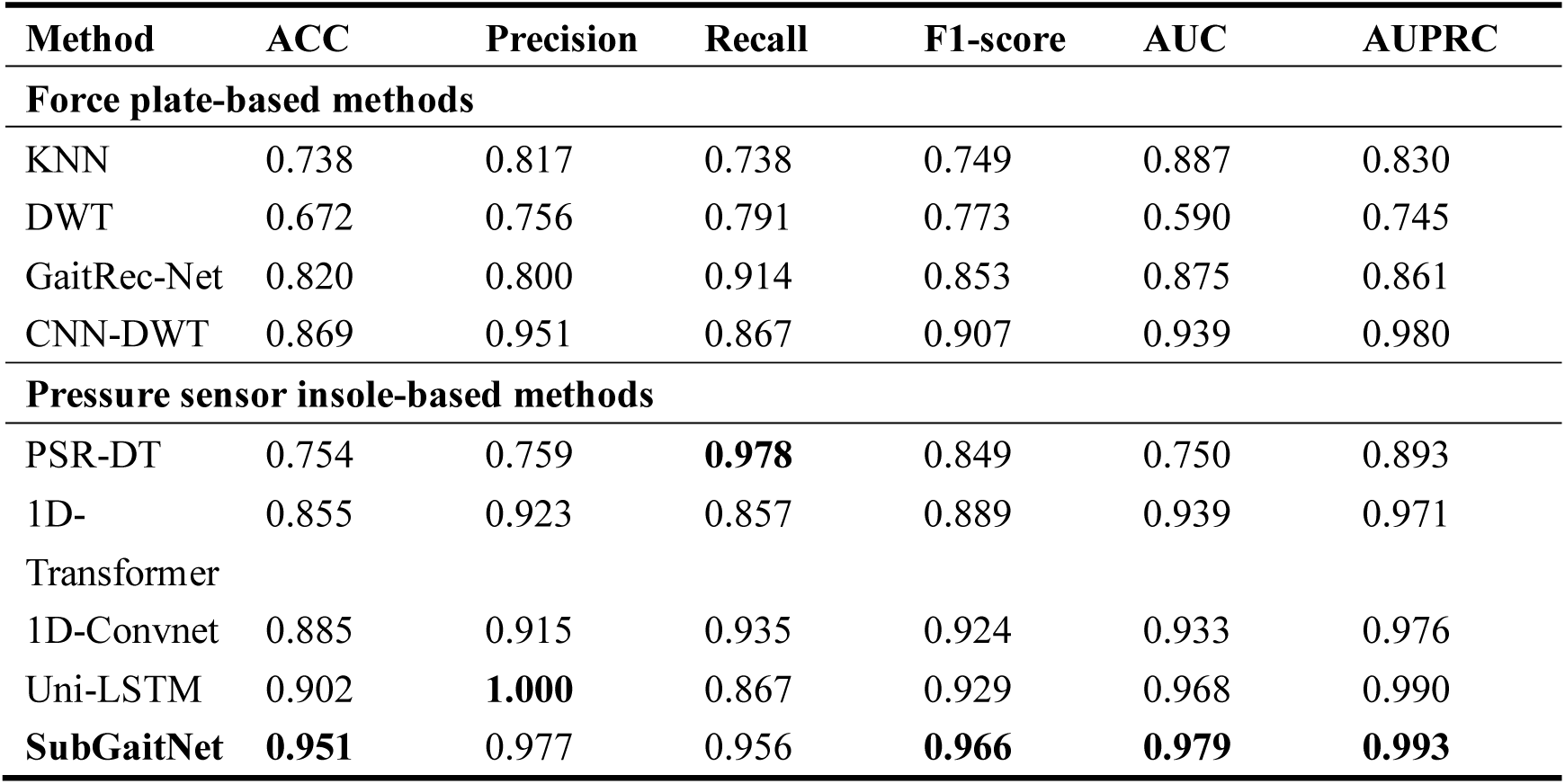
Comparison with existing methods for PD screening on the Gait in Parkinson’s Disease dataset.

To further evaluate statistical stability, probability reliability, and potential decision-related value for PD screening, we performed bootstrap confidence interval estimation, calibration analysis, and DCA on the same test split used in the primary experiment. Bootstrap analysis showed 95% confidence intervals of 0.885-1.000 for ACC, 0.920-1.000 for F1-score, 0.944-1.000 for AUC, and 0.979-1.000 for AUPRC (Fig. 7a). The calibration analysis yielded a Brier score of 0.059 and an expected calibration error of 0.051, suggesting acceptable agreement between predicted probabilities and observed event frequencies. DCA showed that, when the threshold probability was approximately 0.15 or higher, SubGaitNet provided greater net benefit than treat-all and treat-none reference strategies. These analyses support the potential use of SubGaitNet probabilities as auxiliary screening evidence rather than stand-alone diagnostic decisions.

**Fig. 7.**
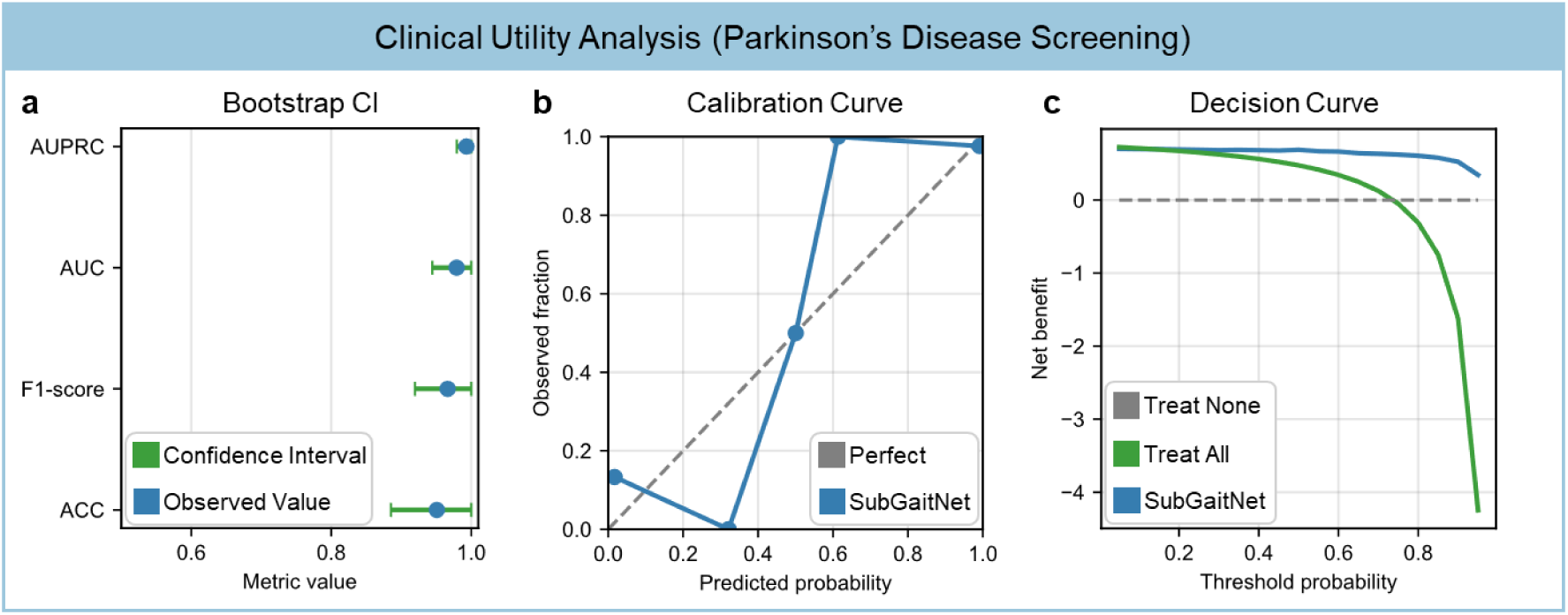
Clinical utility analysis of SubGaitNet for PD screening on the Gait in Parkinson’s Disease dataset.

#### 3.1.2 Fine-grained severity assessment of Parkinson’s disease

In the PD severity (H&Y) assessment task, SubGaitNet showed strong fine-grained classification performance. As illustrated by the confusion matrix in Fig. 8a, most samples were assigned to their corresponding H&Y stages, with the few misclassifications primarily concentrated between clinically adjacent stages (e.g., between H&Y 2.5 and H&Y 2). The multi-class ROC curves (Fig. 8b) suggested discriminative performance across all four severity levels, with AUC values for all classes exceeding 0.956. The t-SNE visualization further showed improved separation among disease stages after feature encoding (Fig. 8c-d), suggesting that the model captured severity-related gait information.

**Fig. 8.**
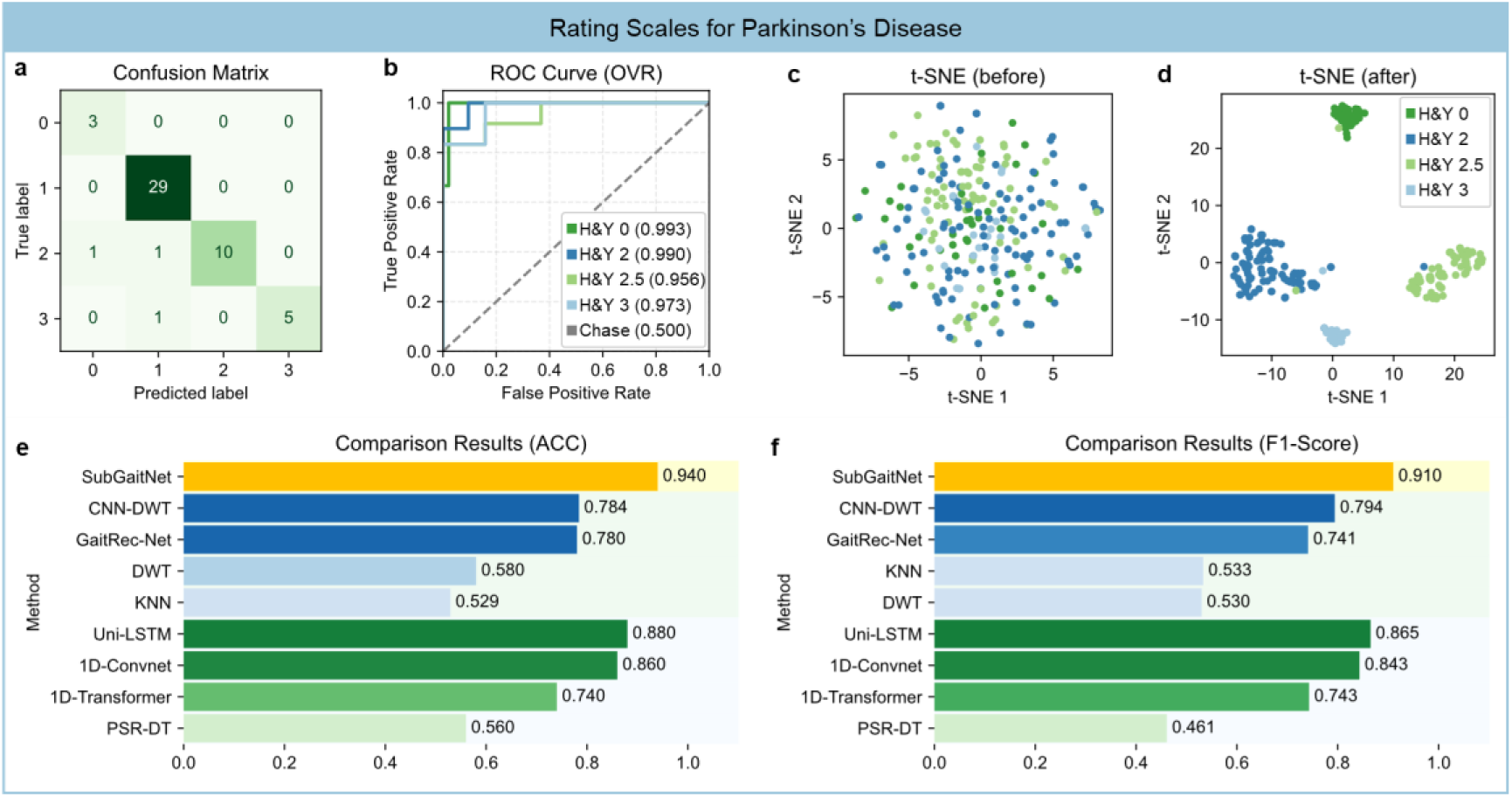
Performance of SubGaitNet for severity staging of PD on the Gait in Parkinson’s Disease dataset.

In comparison with existing methods (Fig. 8e-f; Table 3), SubGaitNet achieved the highest performance, with an ACC of 0.940 and an F1-score of 0.910. Compared with the second-best method (Uni-LSTM, ACC = 0.880), SubGaitNet improved accuracy by 6.0 percentage points. This performance gain supports the added value of phase-aware temporal modeling and explicit gait-variability modeling. Additionally, methods designed for pressure sensor insoles generally outperformed force plate-oriented methods on this dataset, highlighting the importance of matching the representation strategy to the sensing format.

**Table 3.** Comparison with existing methods for severity staging of PD on the Gait in Parkinson’s Disease dataset.

| Method | ACC | Precision | Recall | F1-score | AUC | AUPRC |
| --- | --- | --- | --- | --- | --- | --- |
| **Force plate-based methods** |  |  |  |  |  |  |
| KNN | 0.529 | 0.683 | 0.529 | 0.533 | 0.852 | 0.652 |
| DWT | 0.580 | 0.532 | 0.583 | 0.530 | 0.720 | 0.415 |
| GaitRec-Net | 0.780 | 0.723 | 0.831 | 0.741 | 0.937 | 0.917 |
| CNN-DWT | 0.784 | 0.775 | 0.855 | 0.794 | 0.940 | 0.885 |
| **Pressure sensor insole-based methods** |  |  |  |  |  |  |
| PSR-DT | 0.560 | 0.434 | 0.542 | 0.461 | 0.872 | 0.708 |
| 1D-Transformer | 0.740 | 0.832 | 0.718 | 0.743 | 0.925 | 0.868 |
| 1D-Convnet | 0.860 | 0.857 | 0.834 | 0.843 | 0.939 | 0.874 |
| Uni-LSTM | 0.880 | 0.906 | 0.856 | 0.865 | 0.941 | 0.863 |
| **SubGaitNet** | **0.940** | **0.921** | **0.917** | **0.910** | **0.978** | **0.936** |

To further evaluate the statistical stability of SubGaitNet in H&Y severity grading and its ability to reflect the ordinal structure of disease stages, we performed bootstrap confidence interval estimation and ordinal-aware error analysis on the same test set used in the primary experiment.The test set contained 50 samples. Bootstrap analysis showed 95% confidence intervals of 0.860-1.000 for ACC, 0.768-1.000 for F1-score, 0.943-1.000 for AUC, and 0.802-1.000 for AUPRC (Fig. 9a).

**Fig. 9.**
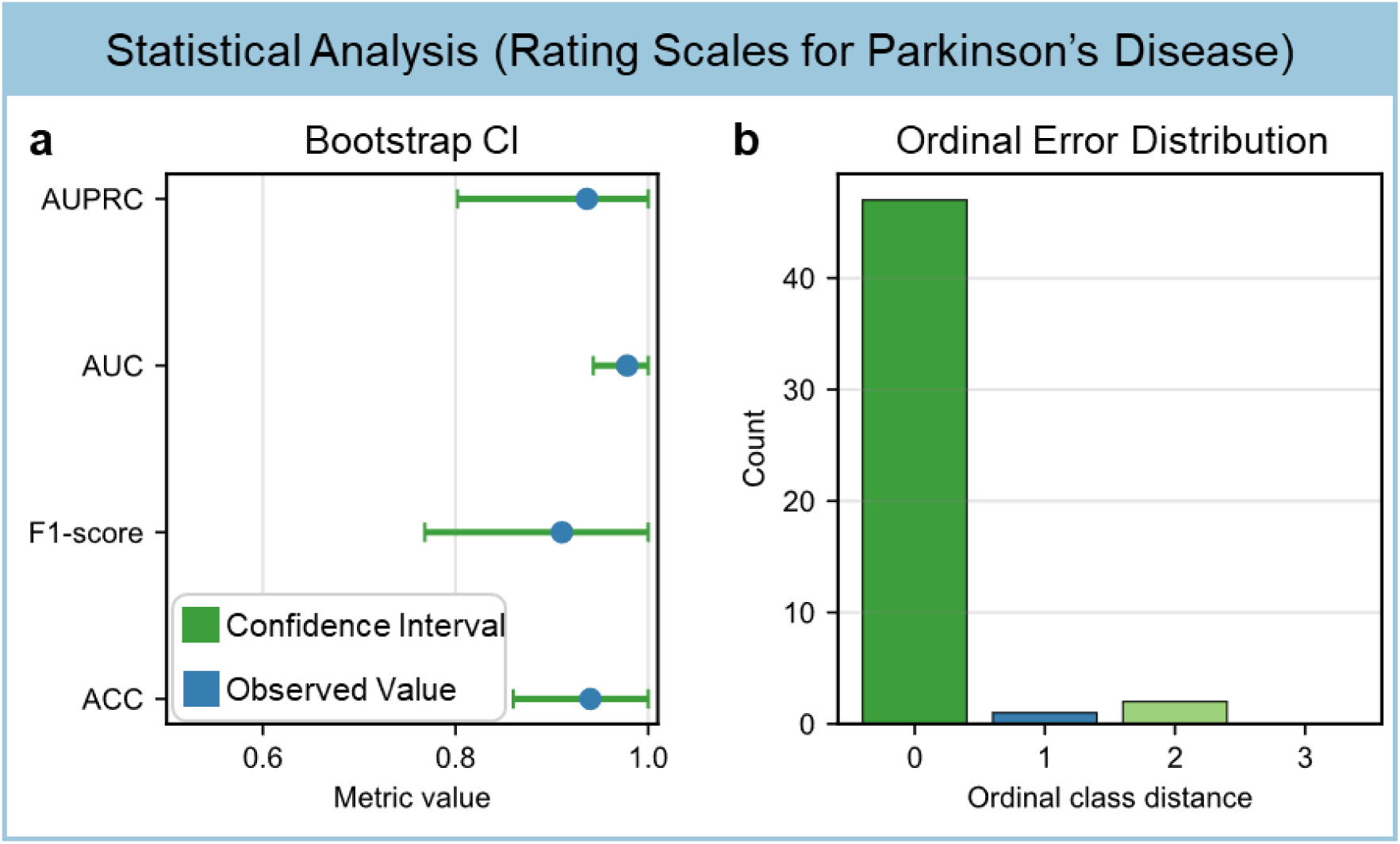
Statistical uncertainty and ordinal error analysis of SubGaitNet for severity staging of PD on the Gait in Parkinson’s Disease dataset.

The quadratic weighted kappa was 0.849, with a 95% confidence interval of 0.618-1.000, suggesting substantial agreement between predicted and reference severity stages. Ordinal error analysis showed that 47 of 50 samples were correctly classified, with an adjacent-stage error rate of 0.020, a non-adjacent-stage error rate of 0.040, and a mean absolute H&Y stage error of 0.080 (Fig. 9b). These results indicate that SubGaitNet not only discriminated among H&Y severity stages but also produced few errors with low ordinal distance, supporting its ability to capture gait representations associated with PD severity progression.

#### 3.1.3 Reliability of wearable gait assessment under sensor failure

In practical clinical applications, pressure sensor insoles frequently experience individual sensor failure due to prolonged usage, which compromises the integrity of the acquired data. As illustrated in Fig. 10a, when a specific sensor fails, the plantar pressure signal (green dashed line) exhibits substantial deviations in amplitude and waveform relative to the original signal (blue solid line), posing a significant challenge to the stability of deep learning models. To assess the effectiveness of SubGaitNet in real-world scenarios, we designed a robustness test simulating sensor failure. In this experiment, the data from individual sensor channels were sequentially zeroed out to emulate sensor malfunctions in practical settings.

**Fig. 10.**
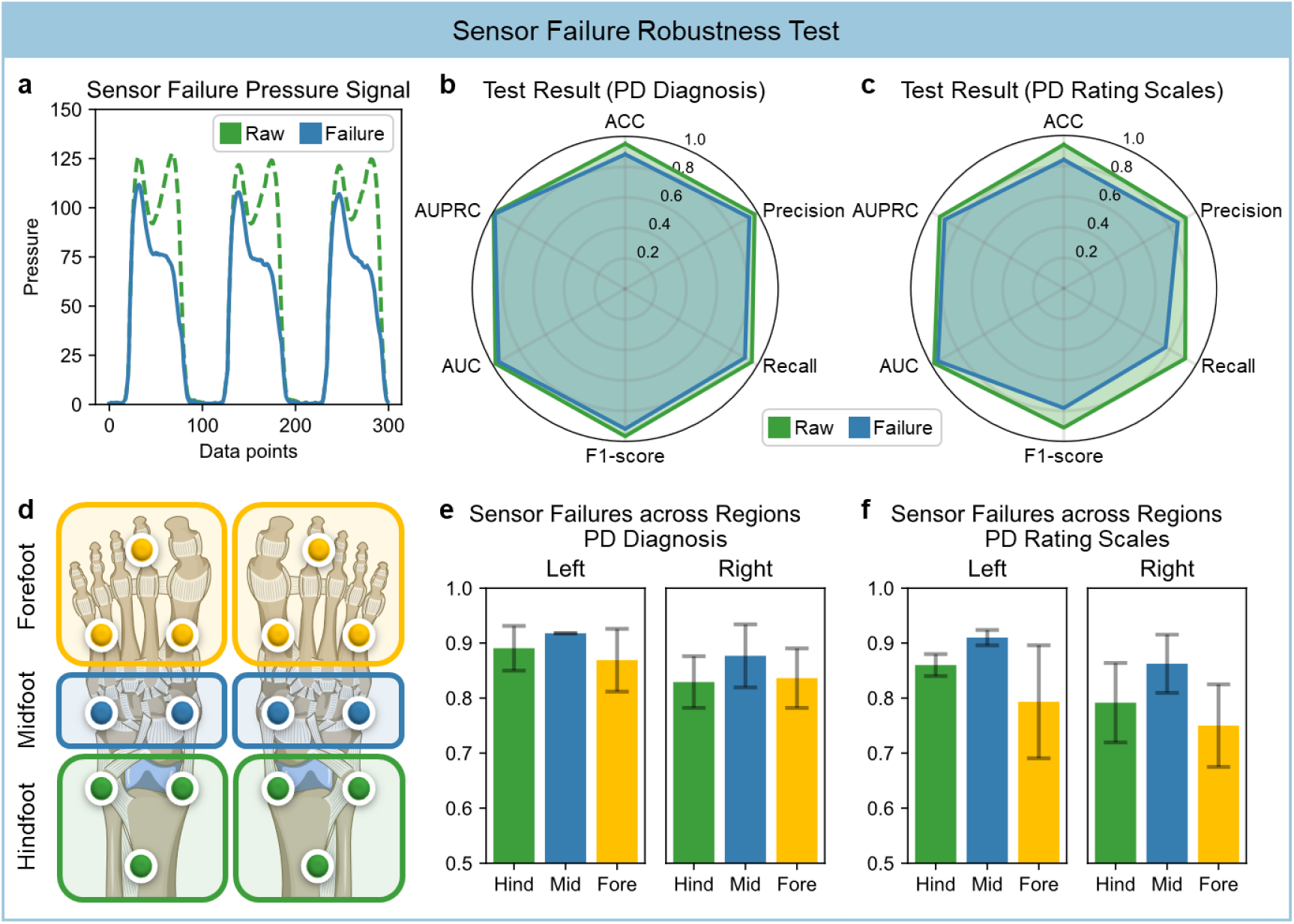
The evaluation results of SubGaitNet under the sensor failure robustness test.

Despite sensor failures, SubGaitNet demonstrated robust performance. Fig. 10b-c present performance comparisons of the model under full-data and sensor-failure conditions in the PD screening and severity grading tasks, respectively. Specifically, in the PD screening task, even with the absence of data from a single sensor, the model achieved an average ACC of 0.882 and an AUC of 0.955, with the F1-score remaining above 0.9. In the more challenging severity grading task, the model remained stable, attaining an average ACC of 0.841 and maintaining an AUC of 0.950. This result indicates that SubGaitNet does not rely on local features from any single sensor, but rather captures global gait coordination patterns. Even with partial information loss, the model utilizes information from the remaining functional channels to sustain high-precision discriminative capabilities.

To further investigate the differential contributions of distinct anatomical plantar regions to the model’s decision-making, we partitioned the plantar sensors into three regions (forefoot, midfoot, and hindfoot; Fig. 10d), and analyzed the specific impact of regional failures on performance. Detailed results are documented in Supplementary Tables 4-5. As shown in Fig. 10e-f, the performance degradation is minimal when midfoot sensors fail. Conversely, the absence of forefoot and hindfoot sensors led to relatively larger performance reductions. This phenomenon is consistent with the pathological characteristics of Parkinsonian gait [23]. The motor impairments in PD patients (e.g., shuffling and festinating gait) are primarily manifested as mechanical abnormalities during the heel-strike (involving the hindfoot) and toe-off (involving the forefoot) phases. Therefore, the forefoot and hindfoot regions convey the most critical pathological discriminative information. This analysis not only verifies the varying importance of different regions but also corroborates that the model has indeed learned biomechanically meaningful key gait features.

#### 3.1.4 Robustness to signal noise perturbations in wearable monitoring

In real-world scenarios, gait signals acquired by wearable devices are frequently corrupted by noise. To evaluate the robustness of SubGaitNet under degraded signal quality, we designed a noise perturbation experiment. Zero-mean Gaussian white noise with varying noise ratios (0.1-0.5) was superimposed on the original plantar pressure signals. As depicted by the signal visualization in Fig. 11a, as the noise ratio increases, the originally smooth pressure waveforms gradually exhibit increasing high-frequency fluctuations. When the noise ratio reaches 0.5, the amplitude of the high-frequency noise approaches that of the underlying signal, substantially masking the original gait cycle features and posing a notable challenge to the model’s robustness.

**Fig. 11.**
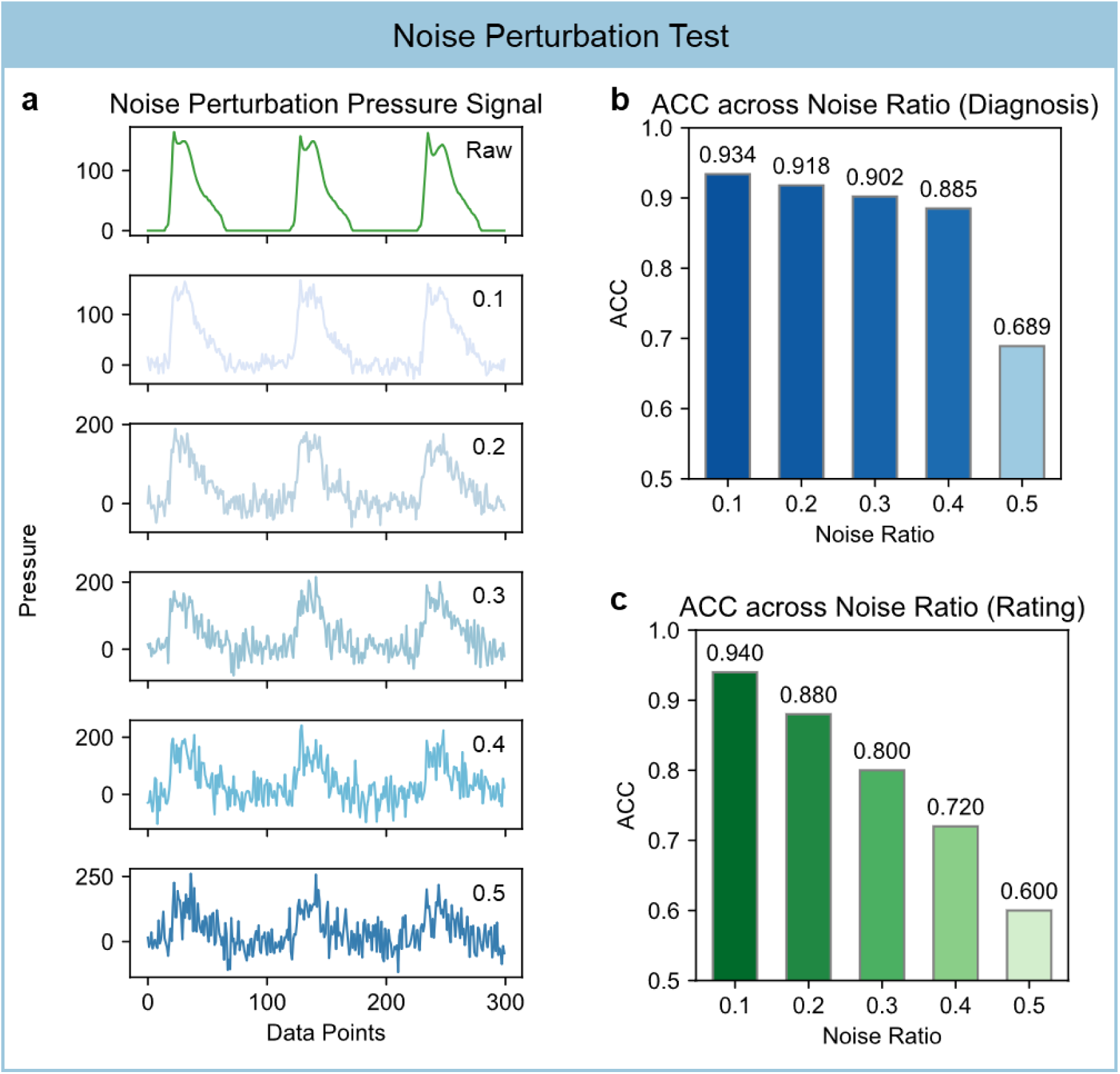
The evaluation results of SubGaitNet under the noise perturbation robustness test.

Fig. 11b-c illustrate the model’s performance trends under varying noise intensities, with detailed data provided in Supplementary Tables 6-7. As the noise ratio increased, screening and severity-assessment performance decreased; however, the model maintained relatively robust performance under low-to-moderate simulated noise. Specifically, in the PD screening task, the model maintained an accuracy above 0.902 when the noise ratio was below 0.3. Even under a 0.4 noise ratio, the accuracy reached 0.885. In the more challenging severity assessment task, the model retained an accuracy of 0.940 at a noise ratio of 0.1. These results suggest that the MS-DRSN module can suppress part of the simulated acquisition noise while preserving gait patterns relevant to assessment.

### 3.2 Force-plate-based musculoskeletal assessment and constrained-input validation

Following the wearable PD assessment scenario, we applied SubGaitNet to a standardized laboratory force-plate setting to evaluate whether the same AI framework could support musculoskeletal gait assessment without architectural redesign. Using the GaitRec dataset, we assessed classification of musculoskeletal impairment sites from 3D GRF and CoP trajectories. We further performed channel-reduction experiments to examine whether clinically useful information could be retained when sensing inputs were simplified.

#### 3.2.1 Musculoskeletal impairment assessment from force-plate gait data

After validating SubGaitNet on pressure sensor insole data, we conducted musculoskeletal impairment assessment experiments on the GaitRec force-plate dataset to evaluate applicability in a standardized gait-laboratory setting. Unlike pressure sensor insoles, which provide vertical pressure signals at distinct plantar locations, force plates acquire GRF signals in three orthogonal directions together with CoP position. As illustrated in Fig. 12a, the task aimed to identify four anatomical impairment locations based on gait patterns: hip, knee, ankle, and calcaneus. We designed two experimental protocols: a four-class task differentiating the four impairment types, and a five-class task that introduced healthy controls to evaluate the model’s ability to distinguish normal from impaired gait while identifying specific impairment sites.

**Fig. 12.**
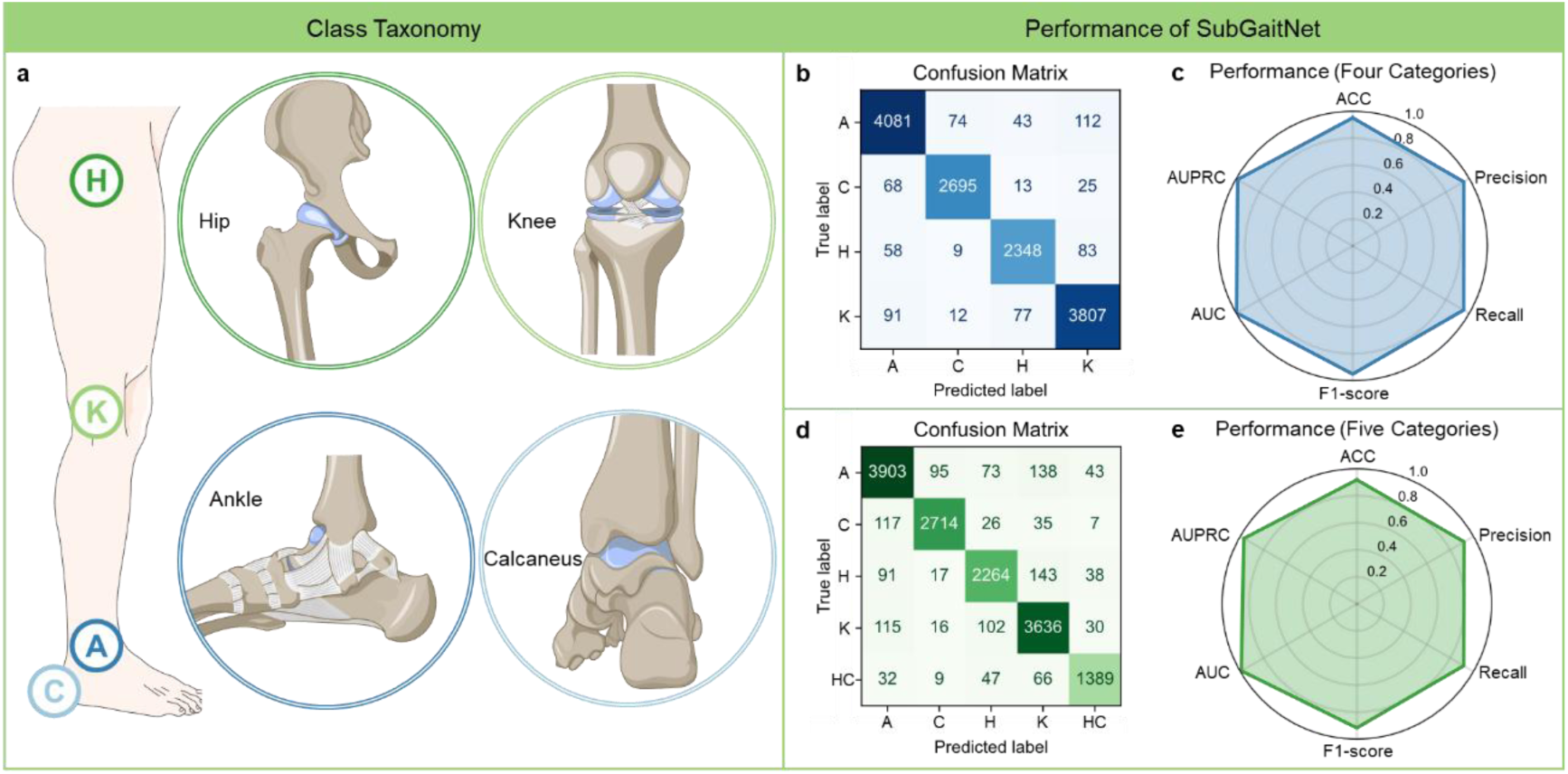
Performance of SubGaitNet for musculoskeletal injuries recognition on the GaitRec dataset.

The experimental results indicate that SubGaitNet also achieved competitive performance on force-plate data. In the four-class task, the model obtained an ACC and F1-score of 0.951. The confusion matrix in Fig. 12b shows that most samples were correctly classified along the diagonal, with limited inter-class misclassification. In the more challenging five-class task, despite the introduction of healthy samples, the model maintained an accuracy of 0.918. Fig. 12d shows that the model separated healthy subjects from subjects with localized musculoskeletal impairments.

To examine the learned feature representation, we used t-SNE to visualize the feature space.

Compared to the raw signals (Fig. 13a, c), the representations extracted by SubGaitNet showed clearer class separation in the embedded space (Fig. 13b, d). This visualization should be interpreted as supportive evidence that the learned representation improves separability, rather than as proof of a causal biomechanical mechanism.

**Fig. 13.**
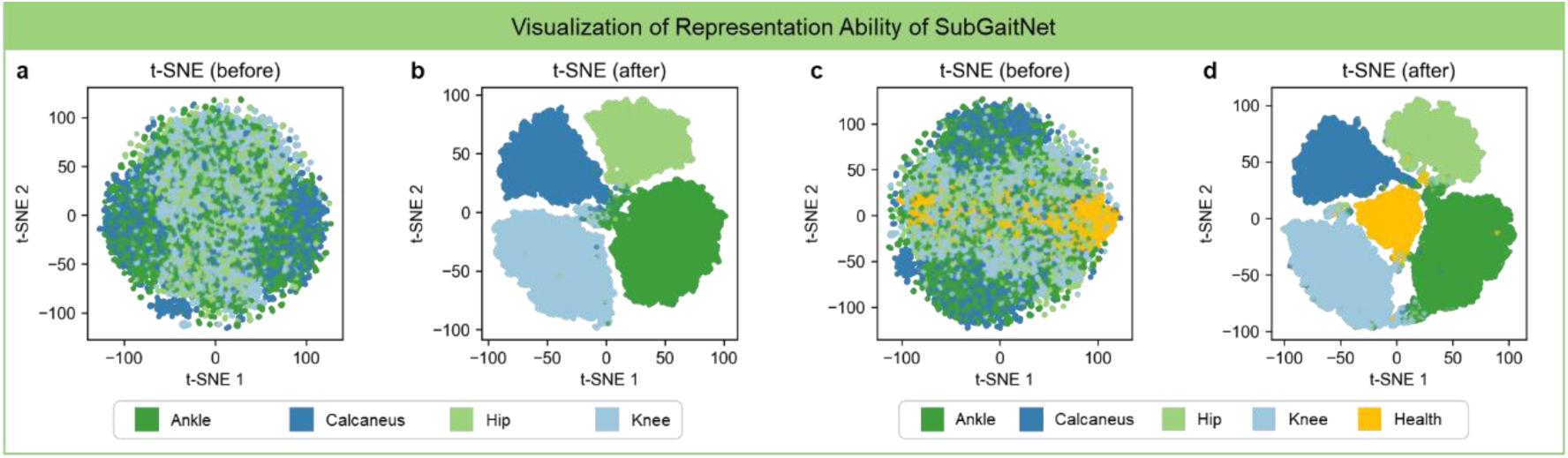
Feature embedding visualization of SubGaitNet for musculoskeletal injuries recognition on the GaitRec dataset.

Fig. 14 and Supplementary Tables 8-9 present the comparative results between SubGaitNet and existing methods. SubGaitNet outperformed the baselines across all evaluation metrics. In the four-class task, SubGaitNet achieved an accuracy of 0.951, representing an improvement of 1.6 percentage points over the second-best method, CNN-DWT (0.935), and 10.2 percentage points over Uni-LSTM (0.849). In the five-class task, the model similarly obtained the highest F1-score (0.917) and AUC (0.990), outperforming both traditional machine learning methods and other deep learning models.

**Fig. 14.**
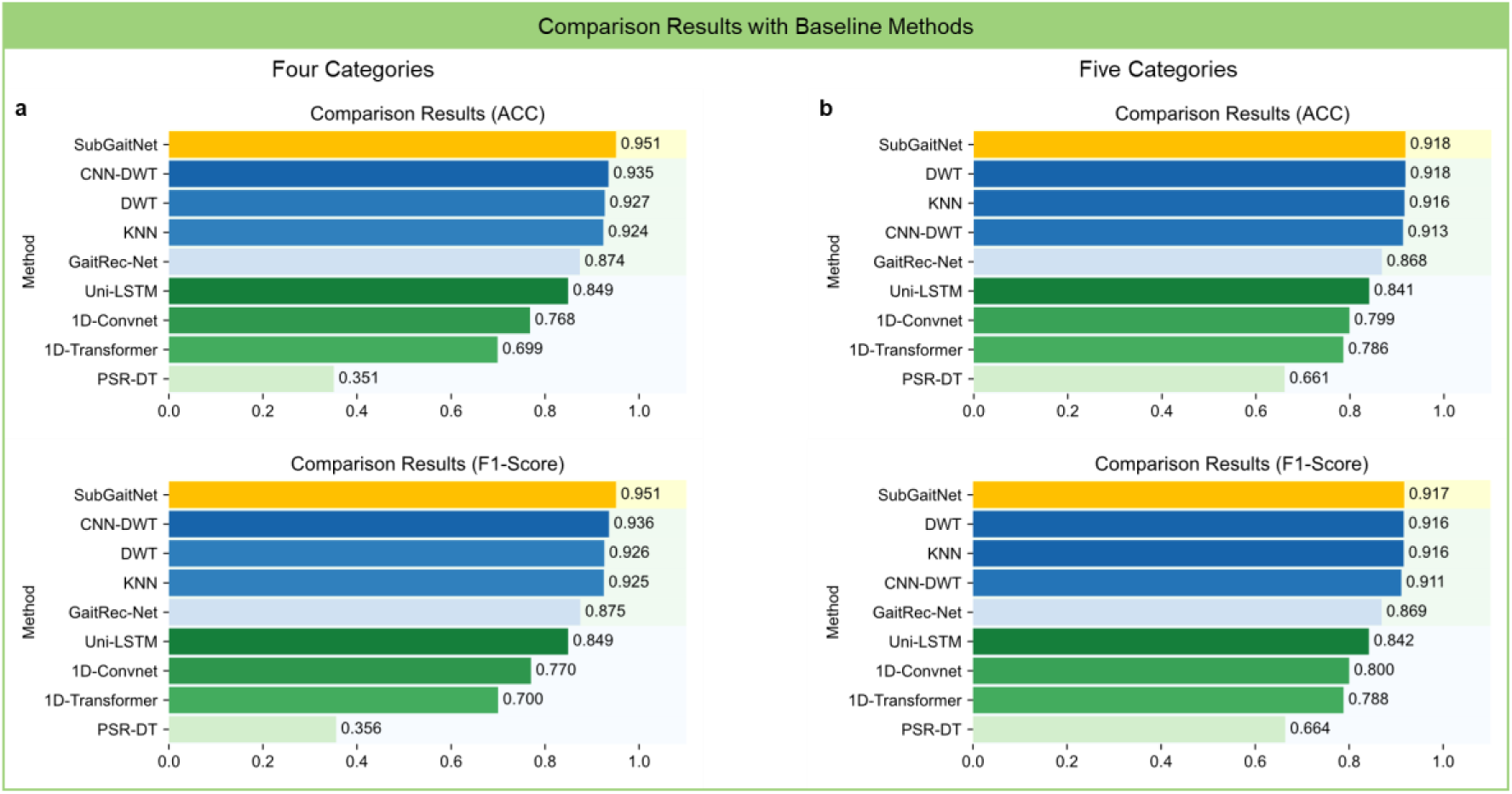
Comparison with existing methods for musculoskeletal injuries recognition on the GaitRec dataset.

Notably, contrary to the results observed on the pressure sensor insole data, methods explicitly designed for force plates (blue) performed better than those designed for pressure sensor insoles (green) on this dataset. This trend may be attributed to the fundamental differences in data characteristics between the two devices: pressure sensor insole data focuses on the localized vertical pressure distribution across the plantar surface, whereas force plate data emphasizes the holistic 3D forces and continuous CoP displacement. Consequently, models tailored to a specific data format may struggle when applied to another sensing format. SubGaitNet achieved the strongest overall performance in both sensing contexts after dataset-specific training, supporting methodological reusability rather than direct device-independent transfer.

To further assess the statistical stability of SubGaitNet for force-plate-based musculoskeletal impairment assessment, we performed bootstrap 95% confidence interval analysis on the GaitRec test sets for both the four-class and five-class tasks. In the four-class task, SubGaitNet achieved 95% confidence intervals of 0.947-0.955 for ACC,0.948-0.955 for F1-score, 0.994-0.995 for AUC, and 0.985-0.988 for AUPRC (Fig. 15a). In the five-class task, where healthy controls were additionally included, the model achieved 95% confidence intervals of 0.914-0.923 for ACC, 0.912-0.922 for F1-score, 0.989-0.991 for AUC, and 0.966-0.972 for AUPRC, respectively (Fig. 15b). These findings indicate that SubGaitNet maintained stable discrimination performanceacross both force-plate-based musculoskeletal assessment tasks, and that the model retained strong overall accuracy and class-separation ability even after the inclusion of healthy controls in the five-class setting.

**Fig. 15.**
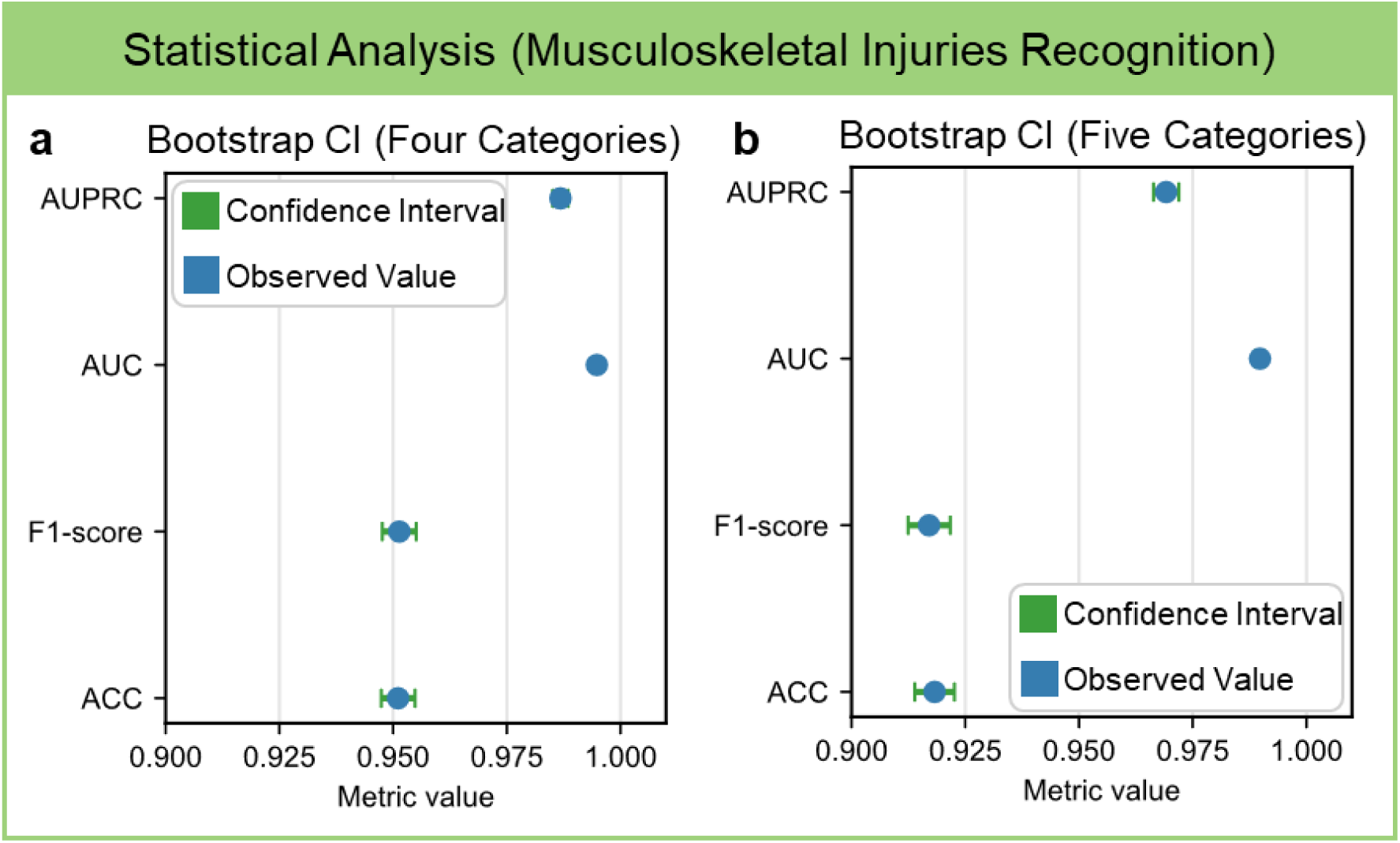
Statistical uncertainty analysis of SubGaitNet for musculoskeletal impairment recognition on the GaitRec dataset.

#### 3.2.2 Constrained-input assessment for accessible gait monitoring

Although force plates provide three-dimensional GRF and CoP data, their high cost may hinder large-scale implementation in practical clinical settings. In contrast, portable and simplified force plates typically capture only the vertical GRF. To evaluate the model’s applicability to low-cost data acquisition devices, we designed a channel reduction experiment. The experiment comprised three input configurations: (i) Raw (5 channels), containing the complete 3D forces and CoP trajectories; (ii) Force Only (3 channels), omitting CoP information to retain only the 3D GRF; and (iii) Vertical Only (1 channel), retaining the vertical GRF. The dataset split remained strictly consistent across all three configurations.

The experimental results are presented in Fig. 16 and Supplementary Tables 10-11. In the absence of CoP data (Force Only), the model’s accuracy in the four-class classification task decreased from 0.951 to 0.896, while the accuracy for the five-class task reached 0.855. Even when relying exclusively on vertical GRF, SubGaitNet maintained an AUC of 0.947 in the four-class task and an AUC of 0.929 in the five-class task. These findings suggest that simplified sensing configurations may retain potentially useful gait assessment information, although validation on actual low-cost devices is needed before clinical deployment.

**Fig. 16.**
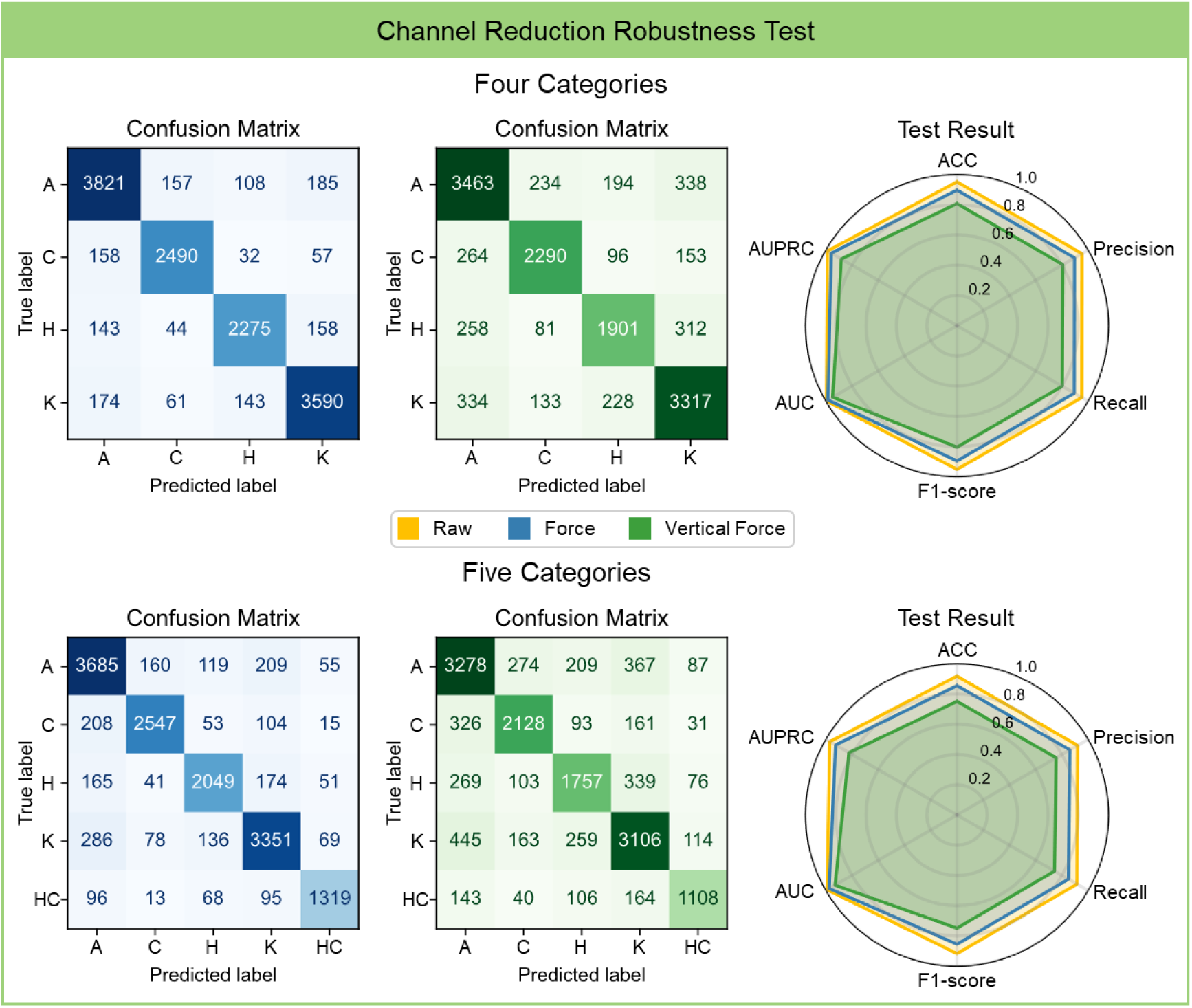
The evaluation results of SubGaitNet under the channel reduction robustness test.

### 3.3 Methodological validity and biomechanical interpretability

To clarify the methodological basis of SubGaitNet and its relevance to medically interpretable AI, we evaluated the contributions of its core components. Parameter sensitivity analyses and ablation studies were used to examine whether temporal slicing, multi-scale residual shrinkage, and dual-branch temporal modeling were necessary for robust gait feature extraction. We then applied SHAP to examine whether the model’s decision evidence aligned with meaningful biomechanical events, such as heel strike and toe-off, rather than reflecting noise or dataset-specific artifacts.

#### 3.3.1 Parameters sensitivity analysis and best architecture selection

To determine the optimal parameter configuration for SubGaitNet, a parameter sensitivity analysis was conducted on the PD screening task. This study focused on the impact of parameters across three key dimensions: (i) the size combination of the small kernel and large kernel within the MS-DRSN module, aiming to balance the receptive fields for local details and global context; (ii) the number of layers and attention heads in the Transformer encoder, to identify the optimal depth for capturing temporal dependencies; and (iii) the effect of the GRF frame length on the completeness of the gait cycle. By employing a controlled-variable approach, we progressively determined the optimal configuration. Detailed results are provided in Supplementary Tables 12-14.

Supplementary Table 12 and Fig. 17a illustrate the model’s performance under various kernel size combinations. The results indicate that the selection of the receptive field is critical for feature extraction. When the large kernel was fixed at 17 × 17, increasing the small kernel size from 3 × 3 to 5 × 5 improved the ACC from 0.918 to 0.951, suggesting that an appropriate expansion of the local receptive field may help capture subtle gait variations. However, a further increase to 7 × 7 resulted in a performance decline (0.934). Ultimately, the combination of Small Kernel = 5 × 5 and Large Kernel = 17 × 17 achieved an optimal balance between local features and long-range dependencies, yielding the highest ACC of 0.951.

**Fig. 17.**
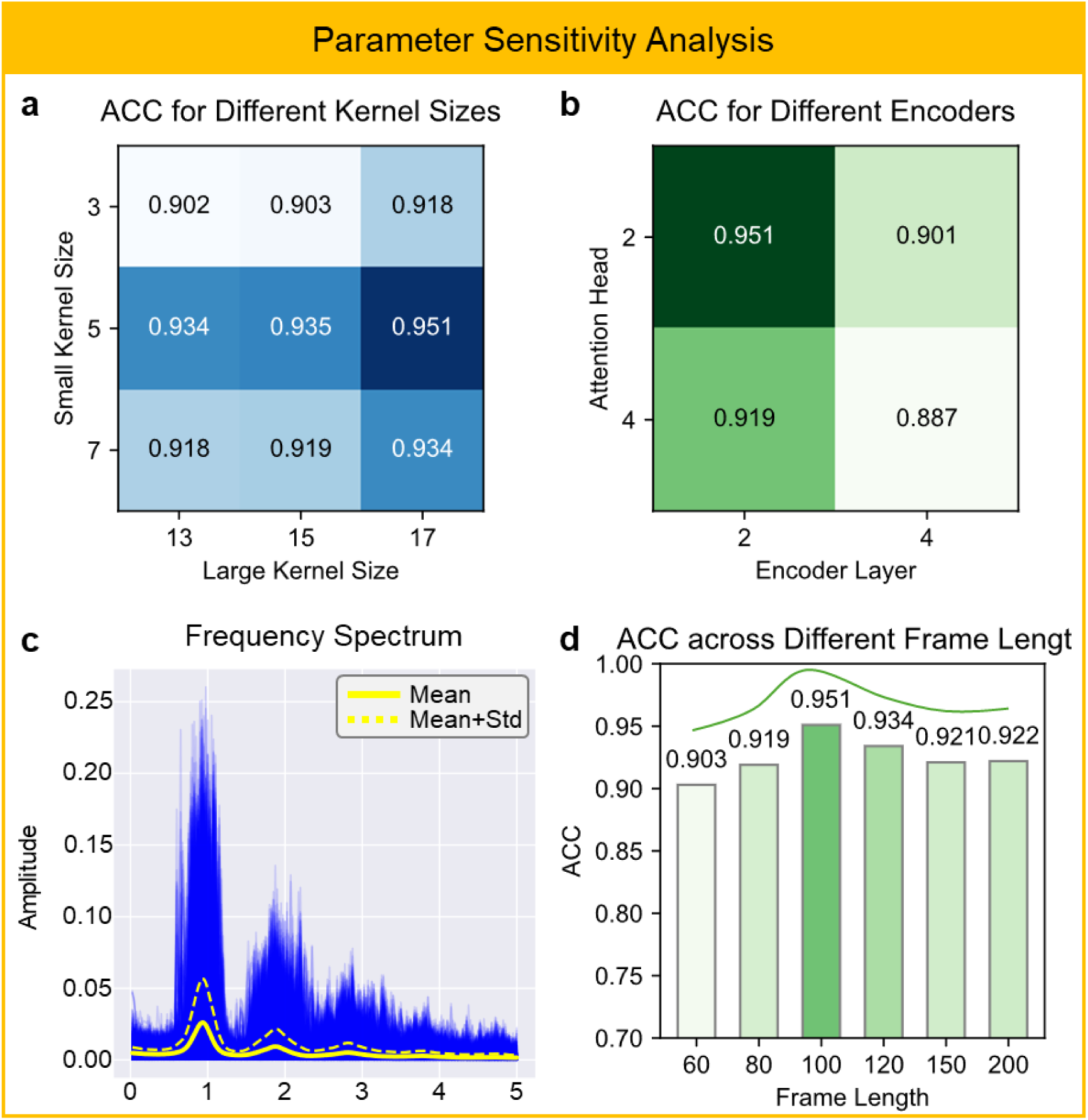
The analysis results of parameters sensitivity.

Following the determination of the convolutional structure, we analyzed the structural parameters of the Transformer. As shown in Supplementary Table 13 and Fig. 17b, shallower network architectures performed better for physiological time-series signals such as GRF. The model achieved the best performance (ACC = 0.951, AUC = 0.979) when configured with 2 encoder layers and 2 attention heads. Conversely, increasing the network depth and width (e.g., to 4 layers or 4 heads) led to a performance decrease (ACC dropped to 0.887). Therefore, the kernel combination of (5 × 5, 17 × 17) and the Transformer structure of (2, 2) were established as the standard configuration for SubGaitNet.

The determination of the GRF frame length was based on the physiological periodicity of human gait. As depicted by the spectral analysis in Fig. 17c, the GRF signal exhibits an amplitude peak near 1 Hz. Given the system’s sampling rate of 100 Hz, this spectral characteristic implies that a time window of approximately 100 data points corresponds to a complete gait cycle. This physiological assumption was empirically validated by the performance trends shown in Fig. 17d and Supplementary Table 14. When the frame length was set to 100 data points, the model reached a peak accuracy of 0.951. Shorter frames (e.g., 60) resulted in a performance drop to 0.903, likely due to the loss of complete periodic patterns. Conversely, excessively long frames (e.g., 150 or 200) also caused performance degradation (ACCs declined to 0.921 and 0.922), potentially introducing irrelevant noise or redundant phases that interfere with temporal modeling. Consequently, 100 was determined as the optimal GRF frame length for capturing complete pathological dynamics.

#### 3.3.2 Quantifying the contribution of key modules via ablation study

To validate the architectural rationale of SubGaitNet and quantify the contributions of its individual modules, we conducted an ablation study on the PD screening task. We constructed four model variants: (i) without Large-Scale Conv: removing the large-kernel branch in the MS-DRSN to retain only local detail features; (ii) without Small-Scale Conv: removing the small-kernel branch in the MS-DRSN to retain only global receptive field features; (iii) without Transformer encoder: removing the Transformer module to rely solely on local convolutions and LSTM for feature aggregation; and (iv) without Sub-LSTM: removing the micro sub-temporal extractor to rely entirely on global temporal modeling.

As presented in Supplementary Table 15, the intact SubGaitNet achieved the best performance across all metrics (ACC = 0.951), whereas the removal of any module resulted in performance degradation. Specifically, omitting the Transformer encoder led to the most substantial performance decrease, with the ACC dropping to 0.820, demonstrating that capturing long-range dependencies within GRF signals is critical for identifying pathological gait. Additionally, removing the Sub-LSTM decreased the accuracy to 0.852, indicating that modeling gait variability is equally indispensable. Regarding spatial features, omitting either the large kernel (0.869) or the small kernel (0.885) caused varying degrees of performance loss, verifying the effectiveness of the multi-scale feature fusion mechanism in balancing global contextual information and local details. In summary, all components contribute positively to the overall performance, corroborating the rationale and necessity of the SubGaitNet architecture.

#### 3.3.3 Clinical interpretability and biomechanical analysis

To improve model interpretability for gait analysis, we employed SHAP to quantify the contributions of sensor channels at different plantar locations. This analysis aimed to identify which plantar regions contribute most to the model’s decisions and whether the resulting attribution patterns align with biomechanical abnormalities reported in PD gait. Specifically, we calculated the mean absolute SHAP value across the 16 channels of the left and right pressure sensor insoles as a global importance metric. Here, L and R denote the left and right foot, respectively; channels 1-3 correspond to the hindfoot, 4-5 to the midfoot, and 6-8 to the forefoot.

As shown in Fig. 18 and Supplementary Table 16, the SHAP attributions are higher in the hindfoot and forefoot than in the midfoot, suggesting biomechanically interpretable importance patterns. In the hindfoot region, R1 (0.0881) and L1 (0.0862) exhibited the highest contributions, indicating that the model relies heavily on pressure pattern differences during the initial contact phase. This is consistent with reported kinetic alterations associated with heel-strike attenuation and shuffling gait in PD [25]. In the forefoot region, R7 (0.0773), R6 (0.0752), and L7 (0.0723) also demonstrated relatively high contributions, suggesting that the model utilizes pressure features during the propulsion phase. This phenomenon corresponds to the reduced push-off and diminished propulsive capacity in PD patients, which generate distinguishable differences in forefoot load distribution [26]. In contrast, the midfoot region showed lower contributions, with L5 (0.0329) and R5 (0.0367) among the lowest, suggesting that midfoot-support signals may contribute less to PD screening in this setting. Overall, this spatial importance distribution reveals a direct correspondence between the model’s decision-making process and the biomechanical characteristics of PD pathological gait.

**Fig. 18.**
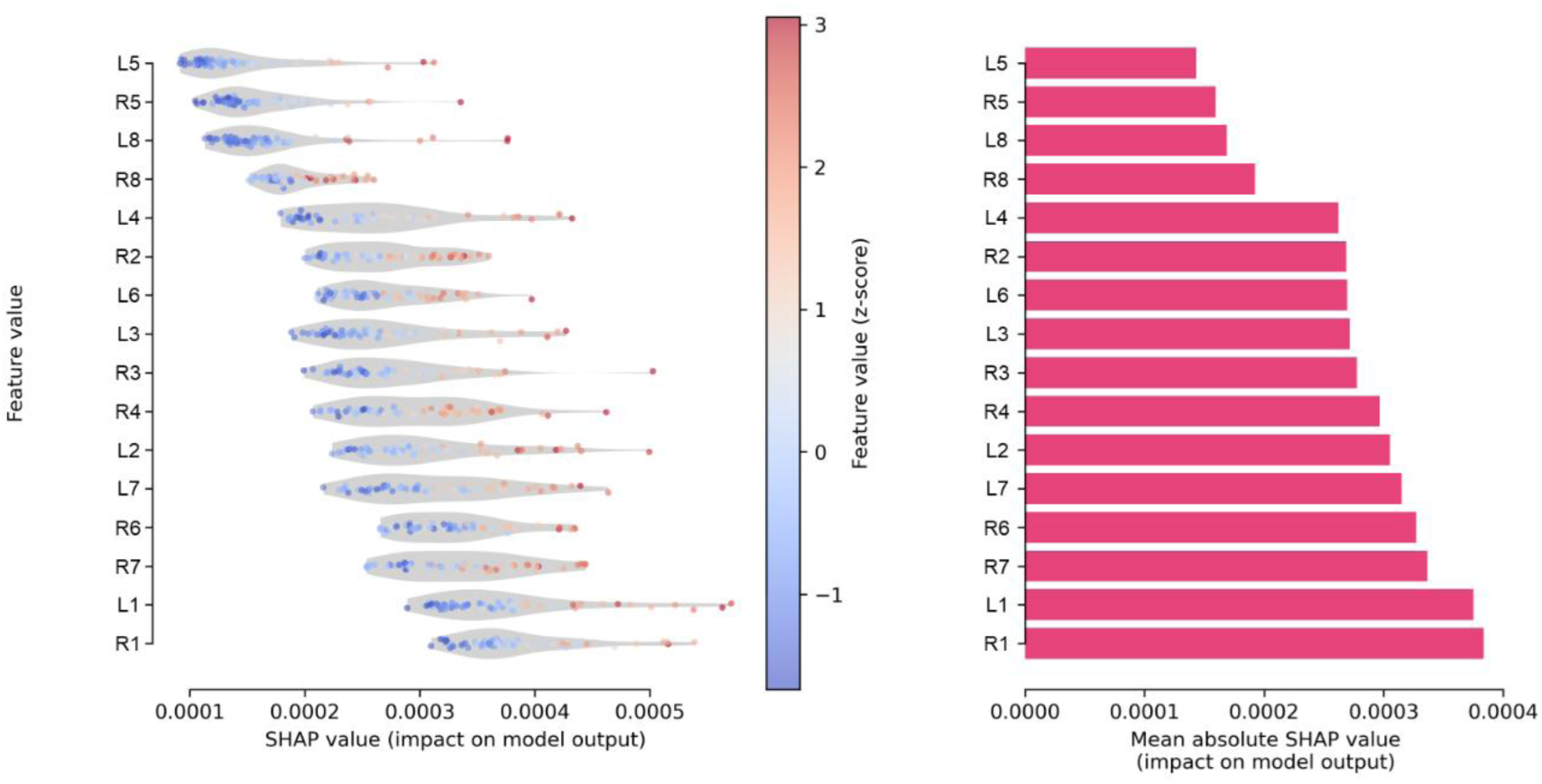
SHAP interpretability analysis of the SubGaitNet.

Furthermore, the SHAP results are consistent with those of the sensor failure robustness test, serving as an explanatory basis for the observed robustness differences. The midfoot channels exhibited lower SHAP contributions; therefore, the model experienced minimal performance degradation when sensors in the midfoot region failed. Conversely, the higher contributions of the hindfoot and forefoot channels corresponded to more pronounced performance drops upon their failure. This consistency not only enhances the credibility of the SHAP explanations but also demonstrates that the discrimination of SubGaitNet does not rely on coincidental noise or irrelevant regions. Instead, it focuses on plantar regions directly associated with abnormal PD gait, thereby supporting the model’s interpretability and reliability in deployment-oriented gait assessment scenarios.

## 4. Discussion

### 4.1 Principal findings and clinical decision-support relevance

This study presents SubGaitNet as a decision-oriented and interpretable AI framework for GRF-based gait health assessment. The work addresses a practical medical AI problem: gait signals contain clinically useful information for neurological and musculoskeletal assessment, but their use in decision-support workflows is limited by heterogeneous sensing devices, variable-length recordings, acquisition noise, sensor failure, and limited access to high-end gait laboratories. By integrating temporal alignment, noise-robust representation learning, long-range gait-phase modeling, and adjacent-frame variability modeling, SubGaitNet supported Parkinsonian gait screening, PD severity assessment, and musculoskeletal impairment classification across two representative GRF sensing contexts.

A key point of this study is that model evaluation was not limited to discrimination accuracy. We further assessed statistical uncertainty, probability calibration, decision-curve net benefit, ordinal error structure for H&Y staging, robustness under sensing uncertainty, constrained-input performance, ablation-based methodological validity, and SHAP-based biomechanical interpretation. These analyses are important for medical AI because gait-based model outputs should be reliable, interpretable, and clinically meaningful enough to support clinician review, rather than functioning as isolated classification scores.

The intended clinical role of SubGaitNet is auxiliary decision support, not autonomous diagnosis. In a potential workflow, GRF data could be acquired using wearable insoles or force-plate systems in community, outpatient, rehabilitation, or home-monitoring settings. The model could then provide screening probabilities, severity-related gait evidence, or impairment-pattern predictions. Abnormal or uncertain outputs would prompt clinician review, and final interpretation would be made together with clinical examination, rating scales, patient history, and follow-up information. This framing keeps the clinical claim appropriately bounded while highlighting the potential value of objective gait-derived evidence for triage, monitoring, and assessment.

### 4.2 Methodological contribution and trustworthy AI evidence

The methodological contribution of SubGaitNet lies in its clinically structured decomposition of GRF gait modeling into four linked representation-learning problems: temporal alignment, denoising, long-range phase-dependency modeling, and adjacent-frame variability modeling. GRF temporal slicing organizes variable-length recordings into gait-phase-related frame sequences; MS-DRSN provides noise-robust local feature extraction; the masked Transformer branch captures long-range gait-phase dependencies while reducing the influence of padded frames; and the Sub-LSTM branch explicitly models adjacent-frame differences that may reflect gait instability and phase-transition irregularity. The ablation results support that these components contribute complementary information rather than serving as arbitrary module stacking.

The use of both wearable-insole and force-plate datasets should be interpreted as a test of methodological reusability, not as evidence for a universal disease-agnostic diagnostic model or direct zero-shot transfer across device families. SubGaitNet retained the same backbone design across representative gait-sensing contexts, but task- and device-specific training remained necessary. This distinction is important because the contribution is a reusable medical AI framework for GRF-based gait assessment, rather than an unsupported claim of immediate cross-device deployment.

Interpretability and robustness further support the trustworthiness of the framework. SHAP analysis indicated that the model emphasized hindfoot and forefoot regions, which are biomechanically consistent with heel-strike and push-off abnormalities relevant to Parkinsonian gait. The overlap between SHAP-highlighted regions and regions associated with larger performance drops in sensor-failure experiments suggests that the learned decision evidence was not concentrated in arbitrary signal components. These findings do not establish causal mechanisms, but they provide clinically understandable evidence that may facilitate clinician review and improve trust in model outputs.

The constrained-input analysis also has practical relevance. Full force-plate systems provide rich 3D GRF and CoP information but may not be available in community clinics or lower-resource rehabilitation settings. The finding that informative performance was retained after removing CoP information, and even when only vertical GRF was used, suggests that simplified sensing configurations may preserve decision-support-relevant gait information. This result should be interpreted as feasibility evidence rather than proof of immediate clinical deployment on low-cost devices.

### 4.3 Limitations and future clinical validation

Several limitations should be acknowledged. First, the current validation is based on public retrospective datasets, and future work should evaluate SubGaitNet in prospective, multi-center cohorts with standardized clinical endpoints, predefined decision thresholds, and clinician-in-the-loop workflows. Second, the current framework requires device-specific retraining and calibration after input-configuration changes; future studies should investigate transfer learning, domain adaptation, calibration drift, and external validation strategies to improve cross-device generalization. Third, although robustness, calibration, decision-curve, and interpretability analyses were included, real-world clinical utility should be further assessed through workflow-based impact studies that quantify whether model outputs improve screening, referral, monitoring, or rehabilitation assessment decisions. Fourth, the current study evaluates auxiliary gait-assessment evidence rather than clinical diagnosis; therefore, claims should remain limited to screening-oriented and assessment-oriented decision support. In summary, SubGaitNet provides a robust and interpretable AI framework for GRF-based gait assessment and may support decision-oriented evaluation in neurological and musculoskeletal care after further prospective validation.

In summary, SubGaitNet provides a reusable, interpretable, and decision-support-oriented AI framework for GRF-based gait assessment across representative neurological and musculoskeletal contexts. Its value lies not only in classification performance, but also in its structured handling of variable-length and noisy gait signals, robustness under clinically plausible sensing uncertainty, and ability to provide biomechanically interpretable decision evidence. Further prospective and workflow-based validation will be required before clinical deployment.

## Code availability

The source code developed for this study, along with the documentation, is available at https://github.com/Ivan020121/SubGaitNet.

## Data availability

The Gait in Parkinson’s Disease dataset used in this study is openly available at https://physionet.org/content/gaitpdb/1.0.0/. The GaitRec dataset used in this study is openly available at https://doi.org/10.6084/m9.figshare.c.4788012.

## Data Availability

https://doi.org/10.6084/m9.figshare.c.4788012

## Acknowledgments

This work was supported by grants from Brain Science and Brain-like Intelligence Technology-National Science and Technology Major Project (No. 2022ZD0209100); Natural Science Foundation of China (no. 82571771) and Natural Science Foundation of Shanghai (no: 25ZR1401167).

## Author contributions: CrediT

Wenhao Li designed and implemented the core methodology, performed the experiments, and wrote the initial draft of the manuscript. Shi Chang conceived the study, supervised the project, contributed to methodology refinement, and revised the manuscript. Liujinxiang Zhu assisted in refining the methodological framework. Yihang Bao was responsible for data collection and preprocessing. Tianqi Liu contributed to the visualization of results. Han Wang and Guan Ning Lin supervised the project design, offered computational resources and guidance, and contributed to manuscript revision.

## References

1 Baker, R., and Hart, H.M.: ‘Measuring walking: a handbook of clinical gait analysis’ (Mac Keith Press London, 2013. 2013)

2 Chau, T.: ‘A review of analytical techniques for gait data. Part 1: fuzzy, statistical and fractal methods’, Gait & posture, 2001, 13, (1), pp. 49–66

3 Beckham, G., Suchomel, T., and Mizuguchi, S.: ‘Force plate use in performance monitoring and sport science testing’, New Studies in Athletics, 2014, 29, (3), pp. 25–37

4 Chen, J.L., Dai, Y.N., Grimaldi, N.S., Lin, J.J., Hu, B.Y., Wu, Y.F., and Gao, S.: ‘Plantar pressure-based insole gait monitoring techniques for diseases monitoring and analysis: a review’, Advanced Materials Technologies, 2022, 7, (1), pp. 2100566

5 Levinger, P., Lai, D.T., Begg, R.K., Webster, K.E., and Feller, J.A.: ‘The application of support vector machines for detecting recovery from knee replacement surgery using spatio-temporal gait parameters’, Gait & posture, 2009, 29, (1), pp. 91–96

6 Alaqtash, M., Sarkodie-Gyan, T., Yu, H., Fuentes, O., Brower, R., and Abdelgawad, A.: ‘Automatic classification of pathological gait patterns using ground reaction forces and machine learning algorithms’, in Editor (Ed.)^(Eds.): ‘Book Automatic classification of pathological gait patterns using ground reaction forces and machine learning algorithms’ (IEEE, 2011, edn.), pp. 453–457

7 Shuzan, M.N.I., Chowdhury, M.E., Reaz, M.B.I., Khandakar, A., Abir, F.F., Faisal, M.A.A., Ali, S.H.M., Bakar, A.A.A., Chowdhury, M.H., and Mahbub, Z.B.: ‘Machine learning-based classification of healthy and impaired gaits using 3D-GRF signals’, Biomedical Signal Processing and Control, 2023, 81, pp. 104448

8 Pandey, C., Roy, D.S., Poonia, R.C., Altameem, A., Nayak, S.R., Verma, A., and Saudagar, A.K.J.: ‘GaitRec-net: a deep neural network for gait disorder detection using ground reaction force’, PPAR research, 2022, 2022, (1), pp. 9355015

9 Ghosh, M., Nandy, A., Patra, B.K., Anitha, R., and Mohanavelu, K.: ‘Multi-modal Deep Neural Features for Classification of Gait Abnormality’, in Editor (Ed.)^(Eds.): ‘Book Multi-modal Deep Neural Features for Classification of Gait Abnormality’ (IEEE, 2024, edn.), pp. 1–6

10 Hausdorff, J.M.: ‘Gait dynamics in Parkinson’s disease: common and distinct behavior among stride length, gait variability, and fractal-like scaling’, Chaos: An Interdisciplinary Journal of Nonlinear Science, 2009, 19, (2)

11 Vaswani, A., Shazeer, N., Parmar, N., Uszkoreit, J., Jones, L., Gomez, A.N., Kaiser, Ł., and Polosukhin, I.: ‘Attention is all you need’, Advances in neural information processing systems, 2017, 30

12 Yu, Y., Si, X., Hu, C., and Zhang, J.: ‘A review of recurrent neural networks: LSTM cells and network architectures’, Neural computation, 2019, 31, (7), pp. 1235–1270

13 Goldberger, A.L., Amaral, L.A., Glass, L., Hausdorff, J.M., Ivanov, P.C., Mark, R.G., Mietus, J.E., Moody, G.B., Peng, C.-K., and Stanley, H.E.: ‘PhysioBank, PhysioToolkit, and PhysioNet: components of a new research resource for complex physiologic signals’, circulation, 2000, 101, (23), pp. e215–e220

14 Horsak, B., Slijepcevic, D., Raberger, A.-M., Schwab, C., Worisch, M., and Zeppelzauer, M.: ‘GaitRec, a large-scale ground reaction force dataset of healthy and impaired gait’, Scientific data, 2020, 7, (1), pp. 143

15 Zhao, M., Zhong, S., Fu, X., Tang, B., and Pecht, M.: ‘Deep residual shrinkage networks for fault diagnosis’, IEEE Transactions on Industrial Informatics, 2019, 16, (7), pp. 4681–4690

16 He, K., Zhang, X., Ren, S., and Sun, J.: ‘Deep residual learning for image recognition’, in Editor (Ed.)^(Eds.): ‘Book Deep residual learning for image recognition’ (2016, edn.), pp. 770–778

17 Hochreiter, S., and Schmidhuber, J.: ‘Long short-term memory’, Neural computation, 1997, 9, (8), pp. 1735–1780

18 Pandey, C., Roy, D.S., Poonia, R.C., Altameem, A., Nayak, S.R., Verma, A., and Saudagar, A.K.J.: ‘GaitRec-Net: A Deep Neural Network for Gait Disorder Detection Using Ground Reaction Force’, PPAR Res, 2022, 2022, (1), pp. 9355015

19 El Maachi, I., Bilodeau, G.-A., and Bouachir, W.: ‘Deep 1D-Convnet for accurate Parkinson disease detection and severity prediction from gait’, Expert Systems with Applications, 2020, 143, pp. 113075

20 Torghabeh, F.A., Modaresnia, Y., and Hosseini, S.A.: ‘An efficient tool for Parkinson’s disease detection and severity grading based on time-frequency and fuzzy features of cumulative gait signals through improved LSTM networks’, Medicine in Novel Technology and Devices, 2024, 22, pp. 100297

21 Nguyen, D.M.D., Miah, M., Bilodeau, G.-A., and Bouachir, W.: ‘Transformers for 1D signals in Parkinson’s disease detection from gait’, in Editor (Ed.)^(Eds.): ‘Book Transformers for 1D signals in Parkinson’s disease detection from gait’ (IEEE, 2022, edn.), pp. 5089–5095

22 Zeng, W., Yuan, C., Wang, Q., Liu, F., and Wang, Y.: ‘Classification of gait patterns between patients with Parkinson’s disease and healthy controls using phase space reconstruction (PSR), empirical mode decomposition (EMD) and neural networks’, Neural Networks, 2019, 111, pp. 64–76

23 Morris, M.E., Iansek, R., Matyas, T.A., and Summers, J.J.: ‘The pathogenesis of gait hypokinesia in Parkinson’s disease’, Brain, 1994, 117, (5), pp. 1169–1181

24 Lundberg, S.M., and Lee, S.-I.: ‘A unified approach to interpreting model predictions’, Advances in neural information processing systems, 2017, 30

25 Kimmeskamp, S., and Hennig, E.M.: ‘Heel to toe motion characteristics in Parkinson patients during free walking’, Clinical biomechanics, 2001, 16, (9), pp. 806–812

26 Sofuwa, O., Nieuwboer, A., Desloovere, K., Willems, A.-M., Chavret, F., and Jonkers, I.: ‘Quantitative gait analysis in Parkinson’s disease: comparison with a healthy control group’, Archives of physical medicine and rehabilitation, 2005, 86, (5), pp. 1007–1013

