## Supplementary Table 1-16 for "SubGaitNet: A Decision-Oriented and Interpretable AI Framework for Robust GRF-Based Gait Assessment in Neurological and Musculoskeletal Care"

Supplementary Information for

SubGaitNet: A Unified Framework for GRF-Based Gait Analysis across Heterogeneous Gait Acquisition Devices

Supplementary Table 1 Demographic and clinical characteristics of the subjects in parkinson’s disease gait dataset

| **Characteristics** | **PD Patients** | **Healthy Controls** | **p-value** |
| --- | --- | --- | --- |
| Age (years) | 66.30 ± 9.50 | 63.66 ± 8.64 | 0.066 |
| Height (cm) | 167.42 ± 8.49 | 168.23 ± 8.52 | 0.543 |
| Weight (kg) | 72.04 ± 12.10 | 72.59 ± 12.35 | 0.774 |
| Gender (Male/Female) | 58 / 35 | 40 / 33 | - |
| Hoehn & Yahr Score | 2.26 ± 0.34 | - | - |

Supplementary Table 2 Demographic of the subjects in GaitRec dataset

| **Category** | **Number** | **Age(yrs.)** | **Body mass (kg)** | **Sex (m/f)** | **Bi-lateral Trials** |
| --- | --- | --- | --- | --- | --- |
| Health control | 211 | 34.7（13.9） | 73.9（15.6） | 104/107 | 7,755 |
| Hip | 450 | 42.6（12.8） | 82.4（15.6） | 373/77 | 12,748 |
| Knee | 625 | 41.6（12.0） | 84.3（18.6） | 426/199 | 19,873 |
| Ankle | 627 | 41.6（11.4） | 87.0（18.0） | 498/129 | 21,386 |
| Calcaneus | 382 | 43.5（10.4） | 84.0（14.5） | 339/43 | 13,970 |
| Total | 2,295 | 41.5（12.1） | 83.6（17.3） | 1,740/555 | 75,732 |

Supplementary Table 3 Parameter settings of the GRF slicing module for different datasets

| **Dataset** | **Gait in Parkinson’s Disease** | **GaitRec** |
| --- | --- | --- |
| Channel type | Pressure sensor | 3-axis GRF and 2-axis CoP |
| Channel count ($C$) | 16 | 5 |
| Normalized length ($L$) | 12000 | 100 |
| GRF frame length（$Ls$） | 100 | 20 |
| GRF frame count（$T$） | 120 | 5 |

Supplementary Table 4 The evaluation results of SubGaitNet for Parkinson’s disease diagnosis under the sensor failure robustness test

| **Channel** | **ACC** | **Precision** | **Recall** | **F1** | **AUC** | **AUPRC** |
| --- | --- | --- | --- | --- | --- | --- |
| L1 | 0.853 | 0.846 | 0.978 | 0.907 | 0.958 | 0.986 |
| L2 | 0.885 | 0.932 | 0.911 | 0.921 | 0.925 | 0.970 |
| L3 | 0.934 | 0.956 | 0.956 | 0.956 | 0.964 | 0.988 |
| L4 | 0.918 | 0.955 | 0.933 | 0.944 | 0.946 | 0.984 |
| L5 | 0.918 | 0.955 | 0.933 | 0.944 | 0.961 | 0.987 |
| L6 | 0.803 | 0.946 | 0.778 | 0.854 | 0.899 | 0.952 |
| L7 | 0.902 | 0.954 | 0.911 | 0.932 | 0.964 | 0.988 |
| L8 | 0.902 | 0.976 | 0.889 | 0.930 | 0.971 | 0.990 |
| R1 | 0.820 | 0.854 | 0.911 | 0.882 | 0.922 | 0.977 |
| R2 | 0.885 | 0.896 | 0.956 | 0.925 | 0.963 | 0.987 |
| R3 | 0.885 | 0.975 | 0.867 | 0.918 | 0.967 | 0.989 |
| R4 | 0.869 | 0.974 | 0.844 | 0.905 | 0.967 | 0.989 |
| R5 | 0.934 | 0.956 | 0.956 | 0.956 | 0.976 | 0.992 |
| R6 | 0.902 | 0.933 | 0.933 | 0.933 | 0.968 | 0.990 |
| R7 | 0.820 | 0.972 | 0.778 | 0.864 | 0.965 | 0.988 |
| R8 | 0.885 | 0.913 | 0.933 | 0.923 | 0.968 | 0.990 |
| Avg | 0.882 | 0.937 | 0.904 | 0.918 | 0.955 | 0.984 |

Supplementary Table 5 The evaluation results of SubGaitNet for severity staging of Parkinson's disease under the sensor failure robustness test

| **Channel** | **ACC** | **Precision** | **Recall** | **F1** | **AUC** | **AUPRC** |
| --- | --- | --- | --- | --- | --- | --- |
| L1 | 0.860 | 0.837 | 0.783 | 0.801 | 0.946 | 0.876 |
| L2 | 0.880 | 0.957 | 0.854 | 0.888 | 0.969 | 0.944 |
| L3 | 0.840 | 0.889 | 0.813 | 0.812 | 0.969 | 0.929 |
| L4 | 0.920 | 0.914 | 0.896 | 0.894 | 0.979 | 0.938 |
| L5 | 0.900 | 0.907 | 0.875 | 0.875 | 0.982 | 0.941 |
| L6 | 0.680 | 0.581 | 0.542 | 0.532 | 0.862 | 0.765 |
| L7 | 0.880 | 0.865 | 0.866 | 0.852 | 0.966 | 0.914 |
| L8 | 0.820 | 0.941 | 0.729 | 0.791 | 0.937 | 0.924 |
| R1 | 0.800 | 0.903 | 0.658 | 0.722 | 0.955 | 0.894 |
| R2 | 0.900 | 0.907 | 0.875 | 0.875 | 0.982 | 0.963 |
| R3 | 0.800 | 0.686 | 0.542 | 0.580 | 0.887 | 0.830 |
| R4 | 0.920 | 0.914 | 0.896 | 0.894 | 0.978 | 0.955 |
| R5 | 0.860 | 0.845 | 0.833 | 0.794 | 0.966 | 0.895 |
| R6 | 0.860 | 0.872 | 0.825 | 0.831 | 0.972 | 0.912 |
| R7 | 0.800 | 0.882 | 0.662 | 0.685 | 0.881 | 0.790 |
| R8 | 0.740 | 0.883 | 0.658 | 0.683 | 0.966 | 0.907 |
| Avg | 0.841 | 0.861 | 0.769 | 0.782 | 0.950 | 0.899 |

Supplementary Table 6 The evaluation results of SubGaitNet for Parkinson’s disease diagnosis under the noise perturbation robustness test

| **Noise ratio** | **ACC** | **Precision** | **Recall** | **F1** | **AUC** | **AUPRC** |
| --- | --- | --- | --- | --- | --- | --- |
| 0.1 | 0.934 | 0.956 | 0.956 | 0.956 | 0.975 | 0.992 |
| 0.2 | 0.918 | 0.976 | 0.911 | 0.943 | 0.961 | 0.988 |
| 0.3 | 0.902 | 0.953 | 0.911 | 0.932 | 0.971 | 0.991 |
| 0.4 | 0.885 | 1.000 | 0.844 | 0.916 | 0.950 | 0.985 |
| 0.5 | 0.689 | 1.000 | 0.578 | 0.732 | 0.922 | 0.975 |

Supplementary Table 7 The evaluation results of SubGaitNet for severity staging of Parkinson's disease under the noise perturbation robustness test

| **Noise ratio** | **ACC** | **Precision** | **Recall** | **F1** | **AUC** | **AUPRC** |
| --- | --- | --- | --- | --- | --- | --- |
| 0.1 | 0.940 | 0.921 | 0.917 | 0.910 | 0.975 | 0.932 |
| 0.2 | 0.880 | 0.880 | 0.792 | 0.824 | 0.964 | 0.924 |
| 0.3 | 0.800 | 0.936 | 0.688 | 0.760 | 0.941 | 0.878 |
| 0.4 | 0.720 | 0.669 | 0.479 | 0.501 | 0.889 | 0.810 |
| 0.5 | 0.600 | 0.398 | 0.292 | 0.257 | 0.823 | 0.672 |

Supplementary Table 8 Comparison with existing methods for musculoskeletal injuries recognition on the GaitRec dataset (four categories)

| **Method** | **ACC** | **Precision** | **Recall** | **F1-score** | **AUC** | **AUPRC** |
| --- | --- | --- | --- | --- | --- | --- |
| **Force plate-based methods** | | | | | | |
| KNN | 0.924 | 0.927 | 0.923 | 0.925 | 0.991 | 0.978 |
| DWT | 0.927 | 0.926 | 0.926 | 0.926 | 0.951 | 0.876 |
| GaitRec-Net | 0.874 | 0.871 | 0.881 | 0.875 | 0.978 | 0.948 |
| CNN-DWT | 0.935 | 0.937 | 0.934 | 0.936 | 0.993 | 0.982 |
| **Pressure sensor insole-based methods** | | | | | | |
| PSR-DT | 0.351 | 0.427 | 0.405 | 0.356 | 0.727 | 0.394 |
| 1D-Transformer | 0.699 | 0.700 | 0.722 | 0.700 | 0.914 | 0.808 |
| 1D-Convnet | 0.768 | 0.775 | 0.770 | 0.770 | 0.940 | 0.862 |
| Uni-LSTM | 0.849 | 0.846 | 0.858 | 0.849 | 0.971 | 0.933 |
| **SubGaitNet** | **0.951** | **0.952** | **0.951** | **0.951** | **0.995** | **0.987** |

Supplementary Table 9 Comparison with existing methods for musculoskeletal injuries recognition on the GaitRec dataset (five categories)

| **Method** | **ACC** | **Precision** | **Recall** | **F1-score** | **AUC** | **AUPRC** |
| --- | --- | --- | --- | --- | --- | --- |
| **Force plate-based methods** | | | | | | |
| KNN | 0.916 | 0.916 | **0.916** | 0.916 | 0.989 | 0.969 |
| DWT | 0.918 | 0.917 | 0.915 | 0.916 | 0.947 | 0.855 |
| GaitRec-Net | 0.868 | 0.863 | 0.878 | 0.869 | 0.982 | 0.946 |
| CNN-DWT | 0.913 | 0.913 | 0.910 | 0.911 | 0.989 | 0.966 |
| **Pressure sensor insole-based methods** | | | | | | |
| PSR-DT | 0.661 | 0.665 | 0.704 | 0.664 | 0.915 | 0.775 |
| 1D-Transformer | 0.786 | 0.788 | 0.789 | 0.788 | 0.955 | 0.875 |
| 1D-Convnet | 0.799 | 0.798 | 0.808 | 0.800 | 0.962 | 0.893 |
| Uni-LSTM | 0.841 | 0.844 | 0.841 | 0.842 | 0.973 | 0.922 |
| **SubGaitNet** | **0.918** | **0.919** | 0.915 | **0.917** | **0.990** | **0.969** |

Supplementary Table 10 The evaluation results of SubGaitNet under the channel reduction robustness test (four categories)

| **Method** | **ACC** | **Precision** | **Recall** | **F1-score** | **AUC** | **AUPRC** |
| --- | --- | --- | --- | --- | --- | --- |
| Force | 0.896 | 0.896 | 0.894 | 0.895 | 0.983 | 0.958 |
| Vertical | 0.807 | 0.807 | 0.802 | 0.804 | 0.947 | 0.881 |
| **SubGaitNet** | **0.951** | **0.952** | **0.951** | **0.951** | **0.995** | **0.987** |

Supplementary Table 11 The evaluation results of SubGaitNet under the channel reduction robustness test (five categories)

| **Method** | **ACC** | **Precision** | **Recall** | **F1-score** | **AUC** | **AUPRC** |
| --- | --- | --- | --- | --- | --- | --- |
| Force | 0.855 | 0.860 | 0.850 | 0.855 | 0.973 | 0.925 |
| Vertical | 0.751 | 0.756 | 0.743 | 0.749 | 0.929 | 0.823 |
| **SubGaitNet** | **0.918** | **0.919** | 0.915 | **0.917** | **0.990** | **0.969** |

Supplementary Table 12 Impact of kernel sizes in MS-DRSN

| **Small** | **Large** | **ACC** | **Precision** | **Recall** | **F1-score** | **AUC** | **AUPRC** |
| --- | --- | --- | --- | --- | --- | --- | --- |
| 3 | 13 | 0.902 | 0.953 | 0.911 | 0.932 | 0.971 | 0.990 |
| 3 | 15 | 0.903 | 0.950 | 0.905 | 0.927 | 0.970 | 0.987 |
| 3 | 17 | 0.918 | 0.935 | 0.956 | 0.945 | 0.961 | 0.987 |
| 5 | 13 | 0.934 | 0.956 | 0.956 | 0.956 | 0.969 | 0.990 |
| 5 | 15 | 0.935 | 0.952 | 0.952 | 0.952 | 0.976 | 0.989 |
| **5** | **17** | **0.951** | **0.977** | 0.956 | **0.966** | **0.979** | **0.993** |
| 7 | 13 | 0.918 | 0.955 | 0.933 | 0.944 | 0.972 | 0.991 |
| 7 | 15 | 0.919 | 0.951 | 0.929 | 0.940 | 0.969 | 0.984 |
| 7 | 17 | 0.934 | 0.918 | **1.000** | 0.957 | 0.974 | 0.990 |

Supplementary Table 13 Impact of Transformer layers and attention heads

| **Head** | **Layer** | **ACC** | **Precision** | **Recall** | **F1-score** | **AUC** | **AUPRC** |
| --- | --- | --- | --- | --- | --- | --- | --- |
| 2 | 2 | **0.951** | **0.977** | 0.956 | **0.966** | **0.979** | **0.993** |
| 2 | 4 | 0.901 | 0.933 | 0.933 | 0.933 | 0.951 | 0.984 |
| 4 | 2 | 0.919 | 0.911 | **0.976** | 0.943 | 0.933 | 0.960 |
| 4 | 4 | 0.887 | 0.907 | 0.929 | 0.918 | 0.924 | 0.962 |

Supplementary Table 14 Impact of GRF frame lengths

| **Length** | **ACC** | **Precision** | **Recall** | **F1-score** | **AUC** | **AUPRC** |
| --- | --- | --- | --- | --- | --- | --- |
| 60 | 0.903 | 0.952 | 0.909 | 0.930 | 0.960 | 0.985 |
| 80 | 0.919 | 0.950 | 0.927 | 0.938 | 0.921 | 0.955 |
| 100 | **0.951** | **0.977** | **0.956** | **0.966** | **0.979** | **0.993** |
| 120 | 0.934 | 0.909 | 1.000 | 0.952 | 0.979 | 0.989 |
| 150 | 0.921 | 0.908 | 0.884 | 0.894 | 0.935 | 0.886 |
| 200 | 0.922 | 0.944 | 0.903 | 0.919 | 0.948 | 0.897 |

Supplementary Table 15 The results of ablation study

| **Method** | **ACC** | **Precision** | **Recall** | **F1-score** | **AUC** | **AUPRC** |
| --- | --- | --- | --- | --- | --- | --- |
| Without Large Scale Conv | 0.869 | 0.951 | 0.867 | 0.907 | 0.939 | 0.980 |
| Without Small Scale Conv | 0.885 | 0.952 | 0.889 | 0.920 | 0.921 | 0.973 |
| Without Transformer Encoder | 0.820 | 0.840 | 0.933 | 0.884 | 0.897 | 0.960 |
| Without Sub-LSTM | 0.852 | 0.923 | 0.857 | 0.889 | 0.949 | 0.977 |
| SubGaitNet | 0.951 | 0.977 | 0.956 | 0.966 | 0.979 | 0.993 |

Supplementary Table 16 SHAP interpretability analysis of the SubGaitNet

| **Channel** | **Mean Absolute SHAP Value** |
| --- | --- |
| L1 | 0.0862 |
| L2 | 0.0701 |
| L3 | 0.0624 |
| L4 | 0.0603 |
| L5 | 0.0329 |
| L6 | 0.0620 |
| L7 | 0.0723 |
| L8 | 0.0388 |
| R1 | 0.0881 |
| R2 | 0.0617 |
| R3 | 0.0637 |
| R4 | 0.0681 |
| R5 | 0.0367 |
| R6 | 0.0752 |
| R7 | 0.0773 |
| R8 | 0.0442 |
